# Explainable death toll motion modeling: COVID-19 narratives and counterfactuals

**DOI:** 10.1101/2020.07.04.20146423

**Authors:** Adriano Veloso, Nivio Ziviani

## Abstract

Models have gained the spotlight in many discussions surrounding COVID-19. The urgency for timely decisions resulted in a multitude of models as informed policy actions must be made even when so many uncertainties about the pandemic still remain. In this paper, we use machine learning algorithms to build intuitive country-level COVID-19 motion models described by death toll velocity and acceleration. Model explainability techniques provide insightful data-driven narratives about COVID-19 death toll motion models – while velocity is explained by factors that are increasing/reducing death toll pace now, acceleration anticipates the effects of public health measures on slowing the death toll pace. This allows policymakers and epidemiologists to understand factors driving the outbreak and to evaluate the impacts of different public health measures. Finally, our models also predict counterfactuals in order to face the challenge of estimating what is likely to happen as a result of an action.

## Introduction

What are the key factors accelerating the number of deaths due to COVID-19? What is the impact of public health countermeasures such as travel bans and movement restriction? Does mass social isolation effectively help decreasing the death toll rate? What would be the impact on death toll if public health countermeasures are relaxed? Answers to these questions may lead to timely and informed policy decisions, but most of them are still open to debate.

COVID-19 seems to be harder to understand than previously appreciated, and a general law or the complete causal chain behind death toll evolution seem to be unfeasible to obtain in the short term. By contrast, there are multiple competing explanations being reported that attribute different causes to death toll evolution in different countries. While these partial explanations are often guided by our standard understanding of the spread of infectious diseases, the COVID-19 death toll increases and decreases in myriad different ways depending on characteristics and particularities of each country, and there is not a sole cause or even one major explanatory factor.

Here we propose to model the COVID-19 death toll evolution by using the concept of motion in which velocity and acceleration are defined as follows:

- Velocity is equal to the slope of the death toll as a function of time. Thus, velocity is the first order derivative of the number of deaths (deaths*/*day).
- Acceleration is equal to the slope of the velocity as a function of time. Thus, acceleration is the second order derivative of the number of deaths (deaths*/*day^2^).

Figure 1 shows death toll, velocity and acceleration for eight countries: Belgium, Brazil, France, Germany, Italy, Sweden, UK and USA, as of June 5th 2020. Commonly, velocity increases at an alarming exponential rate, but then it follows an approximate bell shaped (normalized Gaussian) curve. The steepness of the curve is determined by how quickly the death toll increases. The peak of velocity coincides with the moment in which acceleration is exactly zero. The minimum acceleration point indicates that the contagion became bounded by the lack of resources [1].

**Fig 1.**
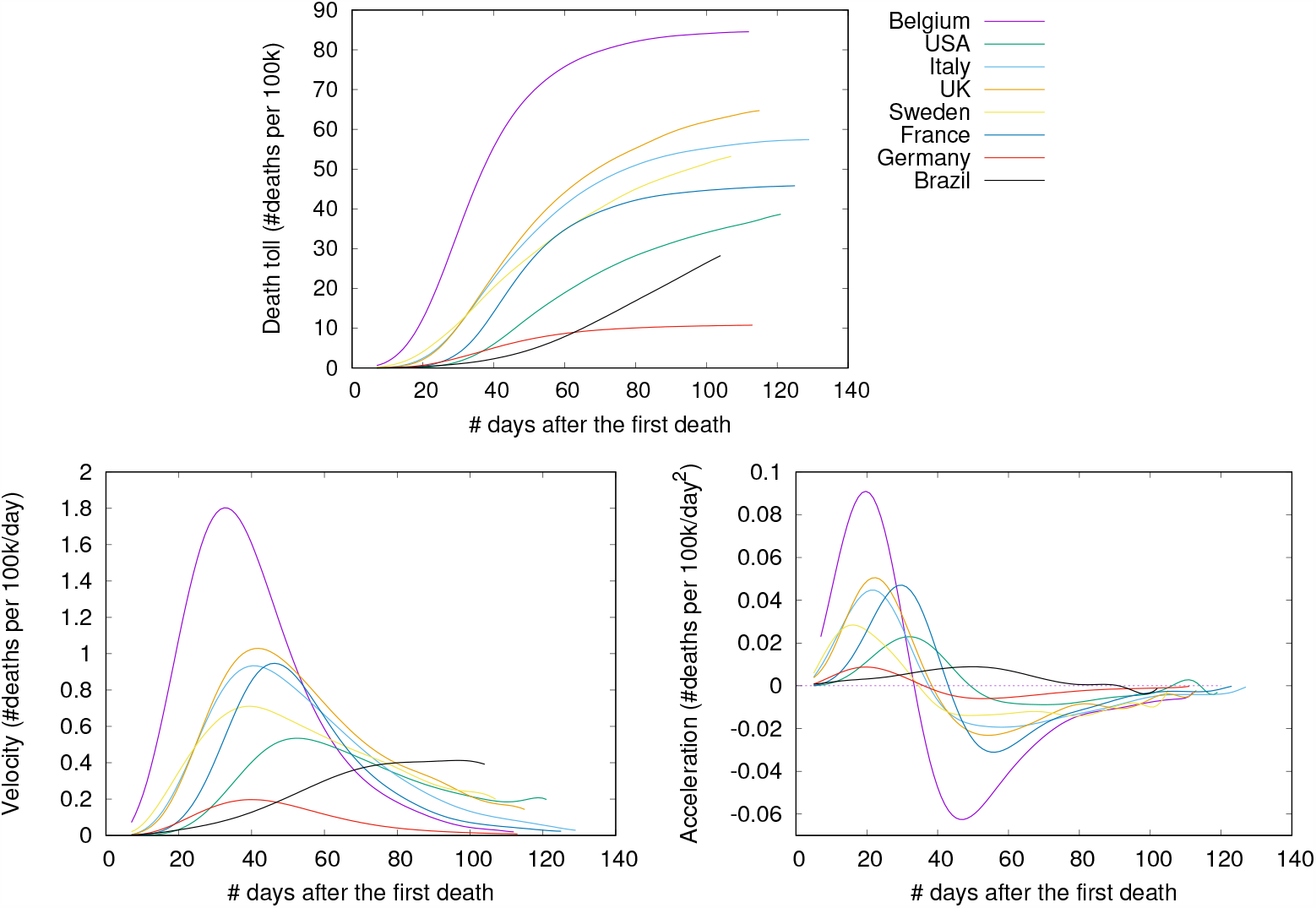
COVID-19 death toll, velocity (first-order derivative) and acceleration (second-order derivative), as of June 5th 2020.

In the figure, curves are aligned by the number of days after the first death reported in the corresponding country. Clearly, COVID-19 death toll evolves differently in each country, with some showing flattened curves but others not. The factors driving the pandemic evolution in each country may vary greatly depending on their specificities and on the countermeasures adopted at each one. By modeling velocity and acceleration across different economy, demography, social, and climate contexts we may unveil non-obvious patterns driving the COVID-19 death toll evolution. Countermeasures (or the lack of them) are often among the factors impacting acceleration the most. Velocity, on the other hand, is mostly impacted by socioeconomic and demographic factors.

In summary, the main contributions of this paper are:

- We model the COVID-19 death toll evolution using the concepts of velocity and acceleration. We build models that predict velocity (deaths*/*day) and acceleration (Δ velocity) to be observed at an arbitrary day of the outbreak. Model predictions come with their explanations, thus providing insightful narratives on why the models behave the way they do. This is an enabling tool for policymakers to understand high-level reasoning behind death toll evolution, so that our models are specially valuable as a planning tool for government officials who need to know about the factors driving the likely trajectory of COVID-19 deaths in their countries. Explanations for acceleration may point to course correction measures that need immediate attention in order to avoid disastrous situations, while explanations for velocity usually point to more intricate factors such as urbanization problems, typical comorbidities, and social inequality issues.
- Our models combine over 200 different factors in order to provide powerful predictions and insightful explanations for death toll evolution. The multitude of factors reflects the interdisciplinary aspects of the pandemic evolution. Clearly, COVID-19 death toll evolution is a complex phenomenon in which factors related to diverse disciplines interact in non-obvious ways, finally resulting in the observed death toll.
- Finally, models are trained on data coming from countries that account for over 99% of all global COVID-19 deaths. These countries are at different outbreak stages, allowing simulations to evaluate potential effects of policy decisions and “what if” reasoning by predicting counterfactuals.

## Materials and methods

In this section we present the data we use in this study and then we discuss how our models were built.

### Data

We employed different sources of data to build our models: (i) the daily COVID-19 death toll curve in each country, (ii) country’s countermeasures in response to COVID-19 pandemics, (iii) community mobility reports, (iv) estimations of critical care beds available for and needed by COVID-19 patients in each country, and (v) country’s development indicators. Next, we present details about the data.

### COVID-19 death toll curve

Data concerning the evolution on the number of deaths comes from case reports provided by the European Center for Disease Prevention and Control (ECDC). The data was frozen on June 5th 2020, comprising 8,926,399 cases and 468,257 deaths reported worldwide. Death reports from 211 countries are presented daily. However, these reports have some issues, including discrepancies in reporting practices across countries. Another issue is the delayed communication of the death, which may result in abrupt variations in consecutive days of the death toll curve of a country. That is, days with under-reported number of deaths are followed by days with over-reported number of deaths. To avoid overfitting the original death toll curve with unnecessarily complex models, we smoothed it using the 7-day rolling average, and from the resulting curve we finally get daily values of velocity (deaths per 100k*/*day) and acceleration (deaths per 100k*/*day^2^).

### Country’s countermeasures in response to COVID-19 pandemics

Measures are implemented to slow the spread of the virus by enforcing physical distance between people. The dates in which these countermeasures were taken in response to COVID-19 pandemics come from the Oxford COVID-19 Government Response Tracker. Data is collected from public sources by a team of Oxford University students and staff from every part of the world [2]. Table 1 shows data about the different measures adopted in response to the COVID-19 pandemics. The value that a countermeasure receives on a specific day is given as the number of days during the outbreak for which it has being taken (or not taken).

**Table 1.**
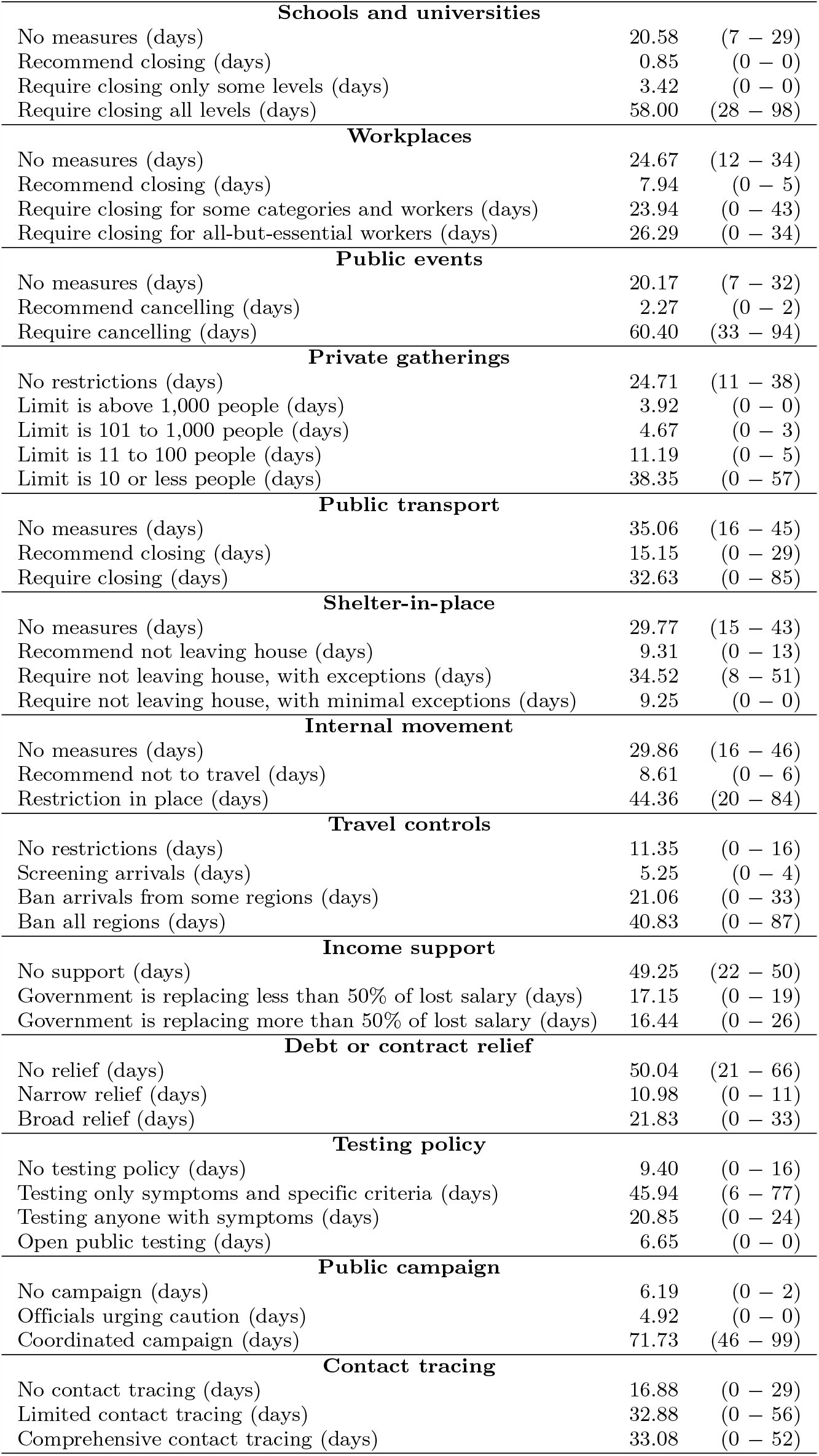
Countermeasures in response to the COVID-19 pandemics. For each countermeasure it is shown, at the peak of velocity, the average followed by first and third quartiles.

| **Schools and universities** |  |  |
| --- | --- | --- |
| No measures (days) | 20.58 | (7 – 29) |
| Recommend closing (days) | 0.85 | (0 – 0) |
| Require closing only some levels (days) | 3.42 | (0 – 0) |
| Require closing all levels (days) | 58.00 | (28 – 98) |
| **Workplaces** |  |  |
| No measures (days) | 24.67 | (12 – 34) |
| Recommend closing (days) | 7.94 | (0 – 5) |
| Require closing for some categories and workers (days) | 23.94 | (0 – 43) |
| Require closing for all-but-essential workers (days) | 26.29 | (0 – 34) |
| **Public events** |  |  |
| No measures (days) | 20.17 | (7 – 32) |
| Recommend cancelling (days) | 2.27 | (0 – 2) |
| Require cancelling (days) | 60.40 | (33 – 94) |
| **Private gatherings** |  |  |
| No restrictions (days) | 24.71 | (11 – 38) |
| Limit is above 1,000 people (days) | 3.92 | (0 – 0) |
| Limit is 101 to 1,000 people (days) | 4.67 | (0 – 3) |
| Limit is 11 to 100 people (days) | 11.19 | (0 – 5) |
| Limit is 10 or less people (days) | 38.35 | (0 – 57) |
| **Public transport** |  |  |
| No measures (days) | 35.06 | (16 – 45) |
| Recommend closing (days) | 15.15 | (0 – 29) |
| Require closing (days) | 32.63 | (0 – 85) |
| **Shelter-in-place** |  |  |
| No measures (days) | 29.77 | (15 – 43) |
| Recommend not leaving house (days) | 9.31 | (0 – 13) |
| Require not leaving house, with exceptions (days) | 34.52 | (8 – 51) |
| Require not leaving house, with minimal exceptions (days) | 9.25 | (0 – 0) |
| **Internal movement** |  |  |
| No measures (days) | 29.86 | (16 – 46) |
| Recommend not to travel (days) | 8.61 | (0 – 6) |
| Restriction in place (days) | 44.36 | (20 – 84) |
| **Travel controls** |  |  |
| No restrictions (days) | 11.35 | (0 – 16) |
| Screening arrivals (days) | 5.25 | (0 – 4) |
| Ban arrivals from some regions (days) | 21.06 | (0 – 33) |
| Ban all regions (days) | 40.83 | (0 – 87) |
| **Income support** |  |  |
| No support (days) | 49.25 | (22 – 50) |
| Government is replacing less than 50% of lost salary (days) | 17.15 | (0 – 19) |
| Government is replacing more than 50% of lost salary (days) | 16.44 | (0 – 26) |
| **Debt or contract relief** |  |  |
| No relief (days) | 50.04 | (21 – 66) |
| Narrow relief (days) | 10.98 | (0 – 11) |
| Broad relief (days) | 21.83 | (0 – 33) |
| **Testing policy** |  |  |
| No testing policy (days) | 9.40 | (0 – 16) |
| Testing only symptoms and specific criteria (days) | 45.94 | (6 – 77) |
| Testing anyone with symptoms (days) | 20.85 | (0 – 24) |
| Open public testing (days) | 6.65 | (0 – 0) |
| **Public campaign** |  |  |
| No campaign (days) | 6.19 | (0 – 2) |
| Officials urging caution (days) | 4.92 | (0 – 0) |
| Coordinated campaign (days) | 71.73 | (46 – 99) |
| **Contact tracing** |  |  |
| No contact tracing (days) | 16.88 | (0 – 29) |
| Limited contact tracing (days) | 32.88 | (0 – 56) |
| Comprehensive contact tracing (days) | 33.08 | (0 – 52) |

To compare the set of measures adopted by each country, we apply the t-Distributed Stochastic Neighbor Embedding (t-SNE) method [3], which embeds the 47 measures present in Table 1 into a 2D space. t-SNE works by minimizing the divergence between two distributions: one that measures pairwise similarities of the input objects and another that measures pairwise similarities of the corresponding low-dimensional points in the embedding. As a result, both *X* and *Y* axes are non-linear combinations of the original 47 dimensions. Finally, the method is applied sequentially for each day of the outbreak, resulting in a 2D trajectory for each country, as shown in Figure 2.

**Fig 2.**
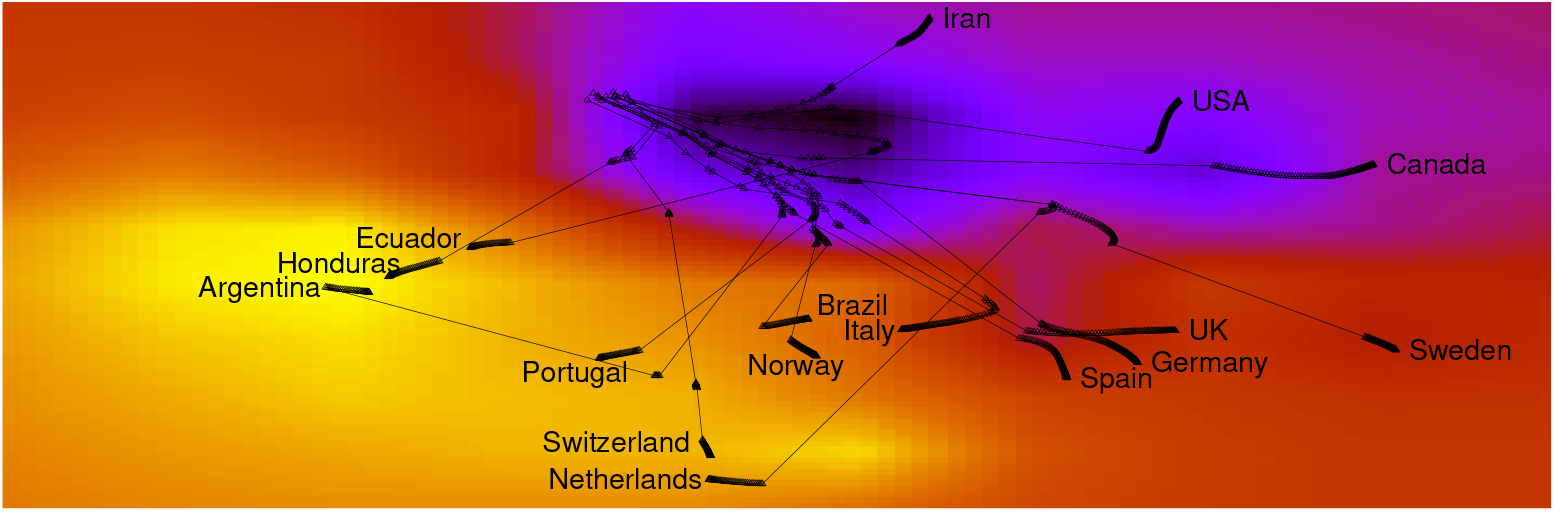
(Color online) Countermeasures adopted by each country forms a trajectory.

In the figure, trajectories are shown over a heatmap composed of clear and dark regions. When the trajectory is passing over a dark region, the corresponding measures are less stringent. By contrast, a trajectory encompasses stringent measures when it passes over a lighter region. Further, the thinner part of a trajectory indicates an abrupt variation on the direction due to changes on the adopted measures. Thicker parts, on the other hand, indicate consecutive days adopting the same set of measures. Finally, countries placed next to each other are currently adopting a similar set of measures. The figure shows a large variation in government responses, with some countries deciding to rise the stringency of response, while others keeping less stringent measures. We follow the definition of stringency presented in [2], that is, the overall stringency level of a country on a given day is calculated by taking the average of the stringencies of the following nine measures: school closing, workplace closing, public events cancelling, gathering size restrictions, public transportation closing, stay at home requirements, restrictions on internal movement, restrictions on international travel, public information campaign. The stringency of each individual measure on a day takes a value between 0 and 100, and thus the overall stringency is given as:

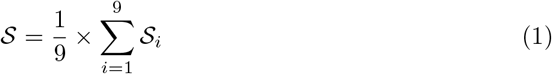

where *𝒮*_*i*_ is the stringency associated with measure *i* on a given day.

### COVID-19 community mobility reports

Human mobility is associated with growth and decline of new cases of COVID-19 [4]. Figure 3 shows how mobility changes as a function of the level of stringency being adopted. As expected, human mobility reduces as stringency increases. The data used to draw the curves in Figure 3 comprises the number of visitors to specific categories of location (e.g., grocery stores, parks, subway and train stations) every day and compares this change relative to a baseline day before the pandemic outbreak. Google Maps^1^ provides anonymized data showing how peoples’ movements have changed throughout the pandemic. Baseline days represent a normal value for that day of the week, given as median value over the five-week period from January 3rd to February 6th 2020, extracted from Google Maps data. The data also comprises the change in average duration spent in residences during the outbreak. We use an offset of three weeks in order to better correlate with the number of deaths by taking into account an approximation of incubation and treatment period lengths.

**Fig 3.**
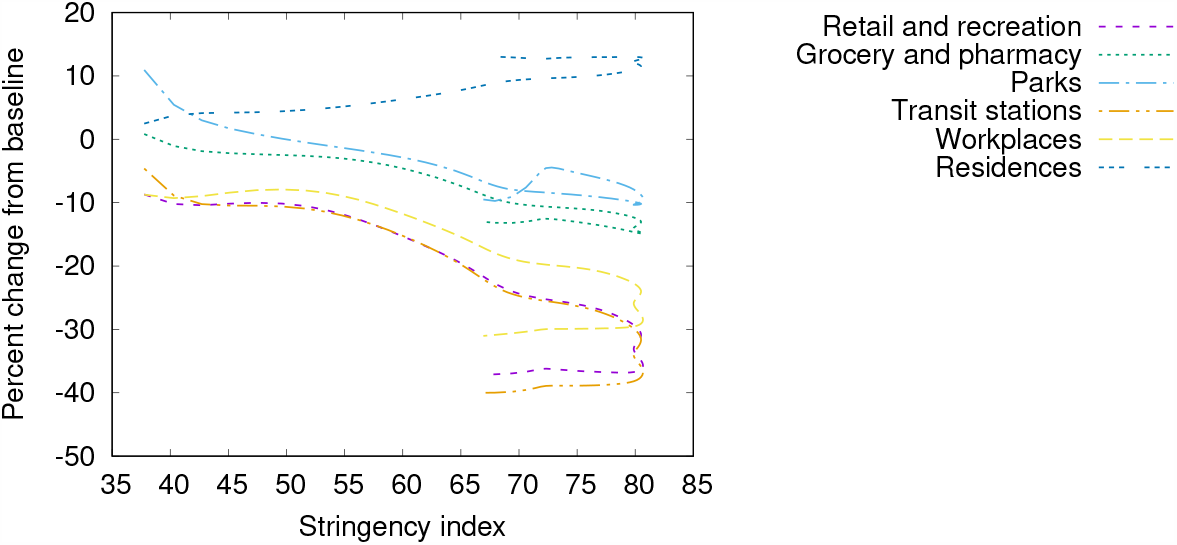
(Color online) Community mobility change.

### Critical care beds available and needed by COVID-19 patients

We use estimations from the Institute for Health Metrics and Evaluation (IHME, University of Washington). These estimations are for COVID-19 patients exclusively, and thus non-COVID patient needs were excluded. Also, the estimation of critical care beds available for COVID-19 patients is obtained by excluding the typical percentage of beds occupied by other patients and emergencies.

### Country’s development data

We use data from the World Bank, which provides a comprehensive dataset on the development of countries around the globe. Table 2 shows a partial list of indicators about health system, climate, socio-economics and demographics for all countries involved in this study. Both temperature and relative humidity are given as the average of the respective values measured in the ten most affected cities in each country, and their values are taken three weeks prior to the current date in order to better correlate with the number of deaths by taking into account an approximation of the incubation and treatment periods.

**Table 2.** Data on health system, climate, socioeconomics and demographics. For each item it is shown the average followed by first and third quartiles.

| Health system preparedness |  |  |
| --- | --- | --- |
| Critical care beds needed/available (on the peak of velocity) | 2.38 | (0.03 – 2.44) |
| Number of nurses (per 100k) | 511.34 | (130.75 – 850.21) |
| Current health expenditure (% of GDP) | 4.19 | (4.01 – 9.44) |
| Climate |  |  |
| Average monthly temperature (C) | 16.83 | (8.72 – 17.37) |
| Average monthly relative humidity (%) | 63.08 | (55.39 – 77.68) |
| Socio-economics |  |  |
| Number of long-term care beds in institutions (per 100k aged 70+) | 2,636 | (1,070 – 6,100) |
| Population living in slums (% urban population) | 21.11 | (4.12 – 35.82) |
| Share of one person households (% households) | 16.56 | (6.48 – 25.87) |
| Income distribution (GINI Index) | 36.93 | (31.50 – 43.70) |
| Demographics |  |  |
| Population in the largest city (% urban population) | 27.85 | (17.14 – 36.71) |
| Population density (inhabitants/km<sup>2</sup>) | 256.52 | (35.73 – 147.85) |
| Urban population (% population) | 65.49 | (53.73 – 83.84) |

Table 3 shows data about age structure, sex-ratio and share of deaths by age. It is worth mentioning that men aged 70 or more correspond to the sub-population at highest risk.

**Table 3.** Data on age structure, sex-ratio by age, and share of deaths by age. For each item it is shown the average followed by first and third quartiles.

| Age structure(% of population) |  |  |
| --- | --- | --- |
| 25+ | 60.72 | (54.53 – 72.70) |
| 50+ | 30.38 | (20.17 – 39.40) |
| 70+ | 7.31 | (3.13 – 11.38) |
| Sex ratio (males per 100 females) |  |  |
| 20–29 | 106.80 | (103.04 – 105.93) |
| 30–39 | 109.70 | (99.20 – 104.55) |
| 40–49 | 108.95 | (96.92 – 103.78) |
| 50–59 | 107.21 | (93.79 – 102.82) |
| 60–69 | 102.44 | (89.01 – 99.95) |
| 70–79 | 89.49 | (81.45 – 94.72) |
| 80–89 | 76.58 | (67.96 – 83.01) |
| 90–99 | 55.51 | (41.78 – 67.14) |
| 100+ | 36.30 | (19.04 – 47.00) |
| Share of deaths by age |  |  |
| 15–49 | 13.92 | (5.27 – 19.62) |
| 50–69 | 25.01 | (19.40 – 29.23) |
| 70+ | 52.33 | (37.03 – 73.04) |

Finally, Table 4 shows a partial list of the risk factors and comorbidities. Our intuition is to correlate COVID-19 death toll motion with an approximation of the size of at-risk group within each country. Prevalence of smoking is based on the population aged 15 years and older who smoke any form of tobacco, including cigarettes, cigars, pipes or any other smoked tobacco products. Obesity is defined as having a body-mass index (BMI) equal to or greater than 30. Diabetes prevalence refers to the percentage of people aged between 20 and 79 years who have type 1 or type 2 diabetes. AIDS prevalence is based on the population aged between 15 and 49 years infected with HIV. Cancer prevalence is based on the population with any form of cancer. Bacilli Calmette-Guérin (BCG) vaccination indicates the countries currently having universal BCG vaccination program. The table also shows death rates from stroke, diabetes, cancer and AIDS, for men and women. Values are standardized so that a value of 1.00 corresponds to the average over all countries we have considered in this study.

**Table 4.** Data on risk factors and comorbidities. For each item it is shown the average followed by first and third quartiles.

| Risk factors and comorbidities: |  |  |
| --- | --- | --- |
| Share of women who smoke (% of women) | 13.61 | (2.00 – 20.00) |
| Share of men who smoke (% of men) | 26.93 | (19.00 – 35.00) |
| Share of women who are obese (% of women) | 24.26 | (19.50 – 28.60) |
| Share of men who are obese (% of men) | 20.24 | (17.10 – 24.20) |
| Diabetes prevalence (% of population) | 7.87 | (5.55 – 8.46) |
| Death rate from cardiovascular disease (per 100k) | 206.20 | (114.77 – 260.94) |
| Share of the population infected with HIV (% of population) | 0.53 | (0.04 – 0.31) |
| Share of population with cancer (% of population) | 1.61 | (0.69 – 2.12) |
| BCG vaccination policy is active | – | 189 (yes) – 22 (no) |
| Death rates by sex (values are standardized): |  |  |
| Death rate from Stroke, female | 0.84 | (0.50 – 1.20) |
| Death rate from Stroke, male | 0.85 | (0.54 – 1.19) |
| Death rate from Cancer, female | 0.97 | (0.94 – 1.13) |
| Death rate from Cancer, male | 0.98 | (0.87 – 1.21) |
| Death rate from Diabetes, female | 0.82 | (0.40 – 1.26) |
| Death rate from Diabetes, male | 0.85 | (0.50 – 1.19) |
| Death rate from AIDS, male | 0.64 | (0.02 – 0.52) |
| Death rate from AIDS, female | 0.58 | (0.01 – 0.33) |

## Models

Our models are particularly useful in situations where many intuitive factors interact in non-obvious ways, which seems to be the case with COVID-19. Specifically, the models are based on two cornerstones:

1. They combine a multitude of factors in order to reflect the interdisciplinary aspects of the pandemic evolution. Model explainability [5] enables inspecting the high-level concepts and reasoning used by our models so that we may formulate possible narratives about the COVID-19 death toll motion defined in terms of velocity and acceleration.
2. They are trained on data from countries that are at different outbreak stages, which enables “what if” reasoning by predicting counterfactuals. This offers powerful simulation possibilities since we may evaluate the potential effects of policies by looking at projections and different scenarios, which is of paramount importance as the pandemic is ongoing.

### Model construction and evaluation

Our models are built using an implementation of the LightGBM algorithm [6]. The LightGBM algorithm produces a complex model composed of hundreds of simple decision trees [7] that are combined into a single model by a process known as boosting [8]. This added complexity allows our models to reach low error levels that are not likely to be obtained by simpler models. Another reason for choosing LightGBM is that it naturally deals with missing values, as some factors are not available for some countries. Basically, LightGBM ignores missing values during a split, and then allocate them to whichever side reduces the loss the most. Models are optimized either to predict the death toll velocity or the death toll acceleration to be observed in the next day. Longer prediction horizons could be achieved by performing next-day predictions in sequence. A model for each country is built and evaluated using discovery and validation datasets, as follows:

- The discovery dataset for a country includes data from all other countries. For the sake of reproducibility: our models were obtained by combining up to 100 simple decision trees with maximum tree-depth of 10, and the learning rate was set to 0.10.
- The validation dataset for a country includes only data from this country. The error of a model with respect to the validation dataset is assessed in terms of the standard Mean Absolute Error (MAE) measure [9], which is given as the mean of absolute values of individual prediction errors on over all instances in the validation dataset. Each prediction error is the difference between observed (e.g., true) and predicted values.

Each instance in both datasets corresponds to a specific day of the outbreak in a country, and it is represented by the factors we present in Tables 1, 2, 3 and 4. Some factors vary daily (i.e., number of critical care beds needed and available), while others remain constant during the entire outbreak period (i.e., population density).

Algorithm 1 describes the main steps of model construction for a specific country. Basically, the algorithm performs an exploratory search for the optimum subset of factors to compose the final model. The exact search approach, however, would require the exhaustive enumeration of all possible combinations of factors, so that one model is obtained for each combination in the power set. Obviously, inspecting all possible subsets of factors is computationally prohibitive. Instead, the algorithm samples the model space by randomly selecting the factors to compose a model. At the end, the best performing models are finally selected.

#### Algorithm 1 Model construction for country *c*

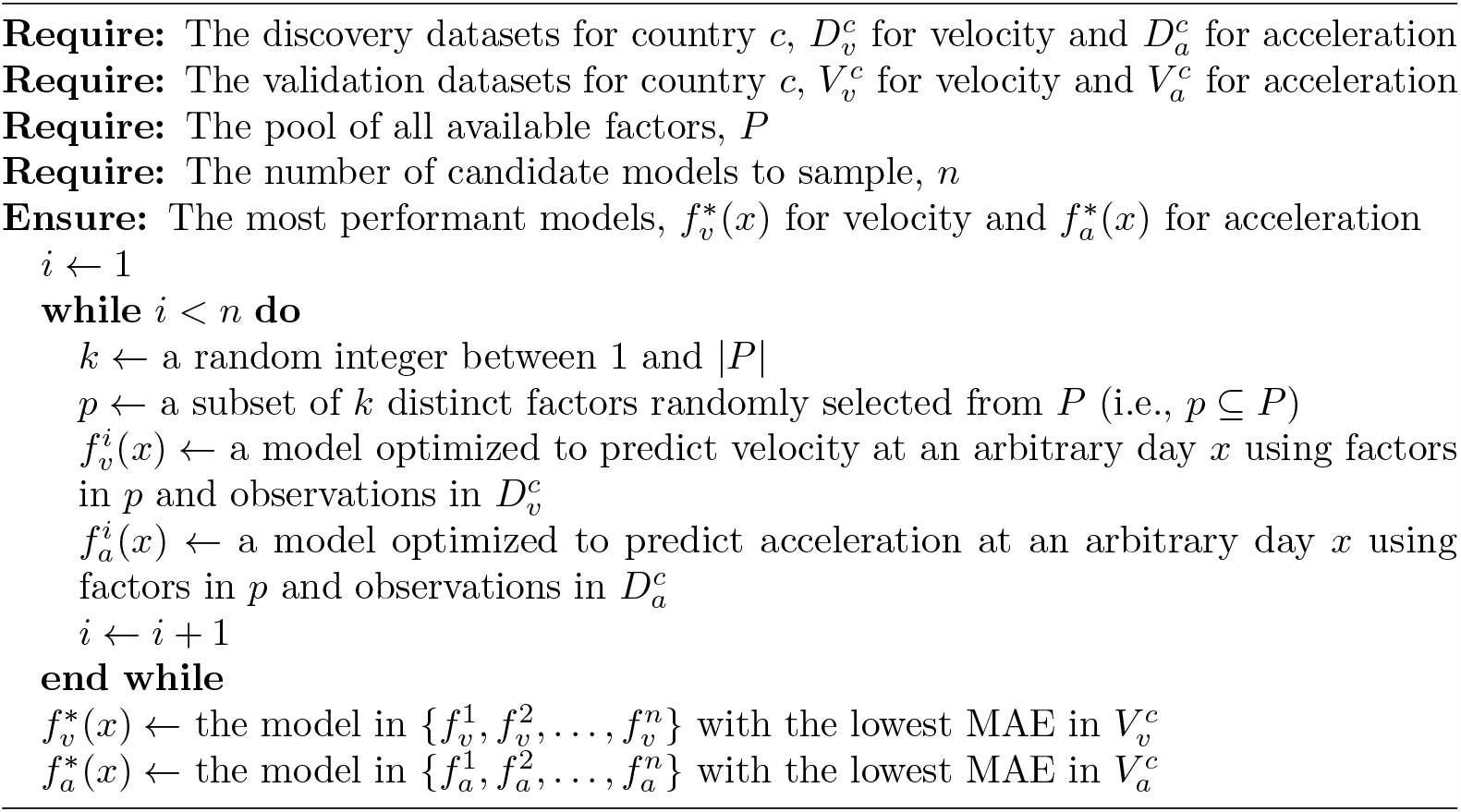

### Model explainability

We assume that a low-error model may be seem as a possible explanation for COVID-19 death toll velocity and acceleration in a country. However, our models are complex as they are obtained from the interplay of a large number of factors. While this provides a more realistic and unified view of COVID-19 death toll motion, understanding how each factor contributes to velocity and acceleration is thus a major challenge. We follow a prominent method that is based on Shapley values from cooperative game theory [10] in order to attribute importance to factors. Specifically, Shapley values can be used to find a fair division scheme that defines how the total importance should be distributed among the factors composing the model [5]. The method is based on the idea that:

i. a specific outbreak day *x* is represented by a set of factors that interact according to a model, thus resulting in the prediction *f* (*x*), and
ii. the magnitude of the difference between *f* (*x*) and the average prediction *E*[*f* (*x*)] (i.e., the average velocity, or the average acceleration in the discovery dataset) is divided among the factors in a unique way that is “fair” given their individual contributions to change the prediction from *E*[*f* (*x*)] to *f* (*x*). Thus, the average contribution each factor has on changing the model prediction is considered to be its importance score.

More formally, the method creates an explanation function *g*(*f, x*) which takes as input a model *f* and the values assumed by the different factors within an outbreak day *x*, and returns importance scores of these factors on day *x*. The importance score associated with a factor can be directly interpreted as the sensitivity that indicates how much the model’s response will vary as the factor has its value increased or decreased.

## Results

In this section, we present results for some countries.^2^ Table 5 shows the Mean Absolute Error (MAE) for velocity and acceleration models of some countries considering the number of days after the first death. In the table, countries are shown in ascending order of error for velocity. The errors of our models are very low, MAEs for prediction of velocity vary from 4.4 × 10^−5^ to 2.4 × 10^−3^ and acceleration from 1.8 × 10^−5^ to 1.9 × 10^−3^, for Pakistan and Sweden, respectively.

**Table 5.**
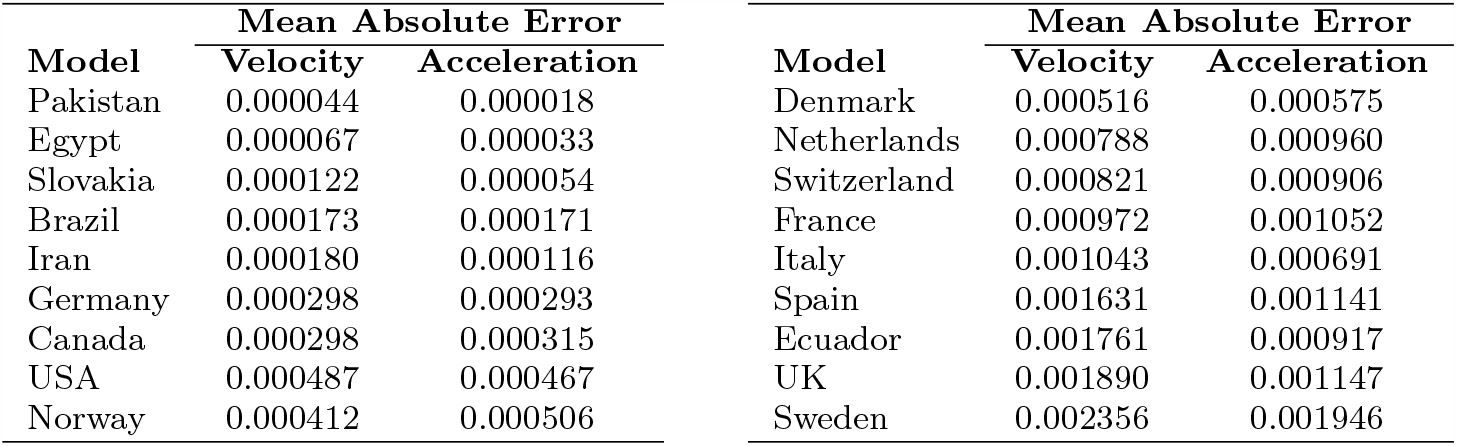
Model effectiveness for some countries. The prediction error is the difference between observed (e.g., true) and predicted values.

| Model | Mean Absolute Error |  | Model | Mean Absolute Error |  |
| --- | --- | --- | --- | --- | --- |
|  | Velocity | Acceleration |  | Velocity | Acceleration |
| Pakistan | 0.000044 | 0.000018 | Denmark | 0.000516 | 0.000575 |
| Egypt | 0.000067 | 0.000033 | Netherlands | 0.000788 | 0.000960 |
| Slovakia | 0.000122 | 0.000054 | Switzerland | 0.000821 | 0.000906 |
| Brazil | 0.000173 | 0.000171 | France | 0.000972 | 0.001052 |
| Iran | 0.000180 | 0.000116 | Italy | 0.001043 | 0.000691 |
| Germany | 0.000298 | 0.000293 | Spain | 0.001631 | 0.001141 |
| Canada | 0.000298 | 0.000315 | Ecuador | 0.001761 | 0.000917 |
| USA | 0.000487 | 0.000467 | UK | 0.001890 | 0.001147 |
| Norway | 0.000412 | 0.000506 | Sweden | 0.002356 | 0.001946 |

For each country we employ model explainability so that we can discuss the main factors driving the predictions for some of these countries. In particular, we employ waterfall plots in order to inspect the main factors explaining velocity and acceleration in important days of the outbreak. Waterfall plots are designed to visually display how the importance of each factor moves the model output from a prior expectation under the discovery dataset distribution to the final model prediction given the evidence of all the factors within the model. Each waterfall plot shows the importance of a set of factors in a given day, so that:

- Factors are sorted from top to bottom by the magnitude of their importance values, with factors of smallest magnitude grouped at the bottom to avoid clutter.
- The importance for each factor is shown as positive (in red) indicating that the factor contributes to increase velocity or acceleration, and negative (in blue) indicating that the factor contributes to decrease velocity or acceleration.

The days between the peak of acceleration and the peak of velocity are of particular importance for evaluating the impact of countermeasures being adopted by public health officials. The days during this period reveal two important aspects: acceleration decreases while velocity keeps increasing, which makes the impact of countermeasures on slowing death toll pace hard to evaluate. However, we are able to present the main factors driving the decrease of acceleration during this period, thus revealing which countermeasures are being effective and also the ones that need further attention.

### Italy, Netherlands and UK - Evaluating countermeasures (and the lack of them)

Figure 4 shows observed and predicted curves and projections for acceleration and velocity for the last fifteen days for Italy. The peak of acceleration in Italy occurred on the 22^*nd*^ day after the first death, while the peak of velocity occurred on the 34^*th*^ day. Figure 5 shows the main factors explaining acceleration on some days within this period. Initially, most of the factors are still contributing to increase acceleration, as expected. Closing schools and universities was the most effective measure at this point. Cancelling public events and the recommendation to close public transportation are still ineffective as they are not yet contributing to decrease acceleration. Five days later, other measures started contributing to decrease acceleration. Finally, at the peak of velocity, most measures are contributing to decrease acceleration, the most effective seem to be self-isolation, closing public transportation, and the cancellation of public events. By contrast, the long time without taking any measure regarding closing schools/universities is now a factor contributing to increase acceleration. Further, some measures may take longer to become effective (i.e., limitation of private gatherings).

**Fig 4.**
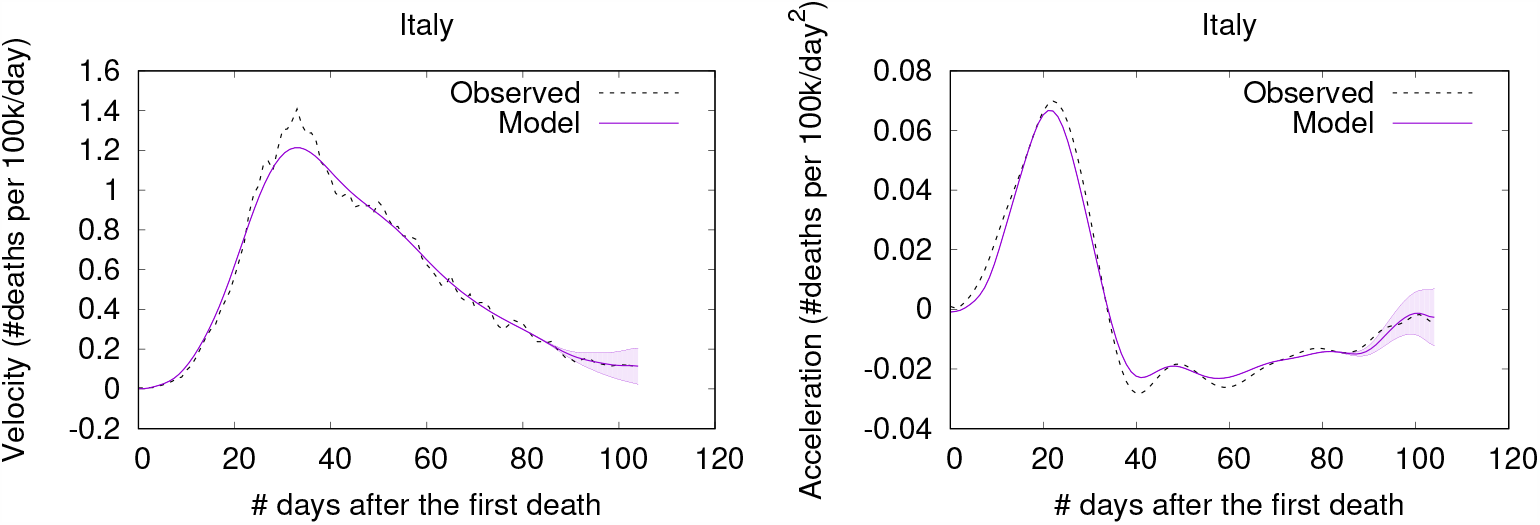
(Color online) Projections for Italy.

**Fig 5.**
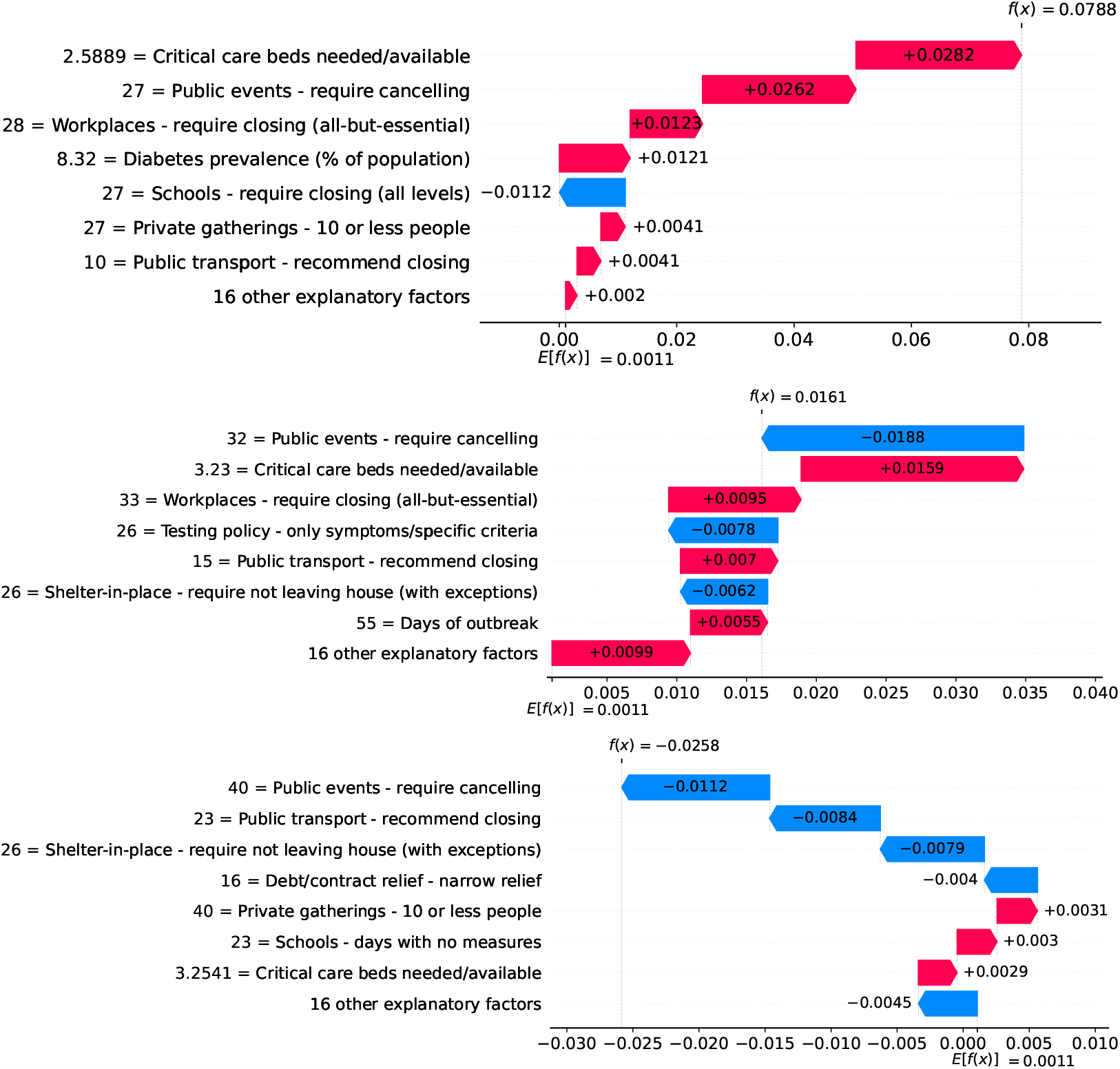
(Color online) Waterfall plots for Italy. Factors driving acceleration on the 22^nd^ (top), 27^*th*^ (middle), and 34^*th*^ day (bottom).

Figure 6 shows observed and predicted curves for Netherlands. The peak of acceleration occurred on the 17^*th*^ day after the first death, while the peak of velocity occurred on the 26^*th*^ day. Figure 7 shows the main factors explaining acceleration on the 20^*th*^ and 26^*th*^ days. Few days after the peak of acceleration (i.e., 20^*th*^ day), measures on restricting internal movement started to decrease acceleration. On the peak of velocity, several factors are contributing to decrease acceleration, but the main ones include cancelling public events and banning arrivals from affected regions.

**Fig 6.**
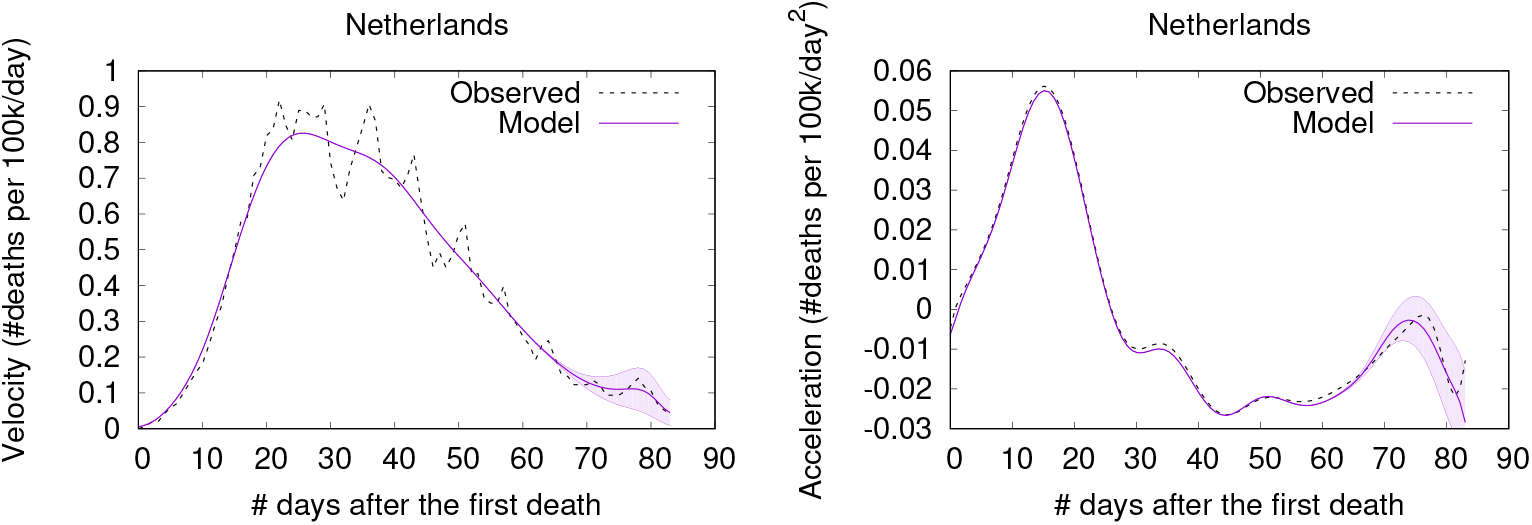
(Color online) Projections for Netherlands.

**Fig 7.**
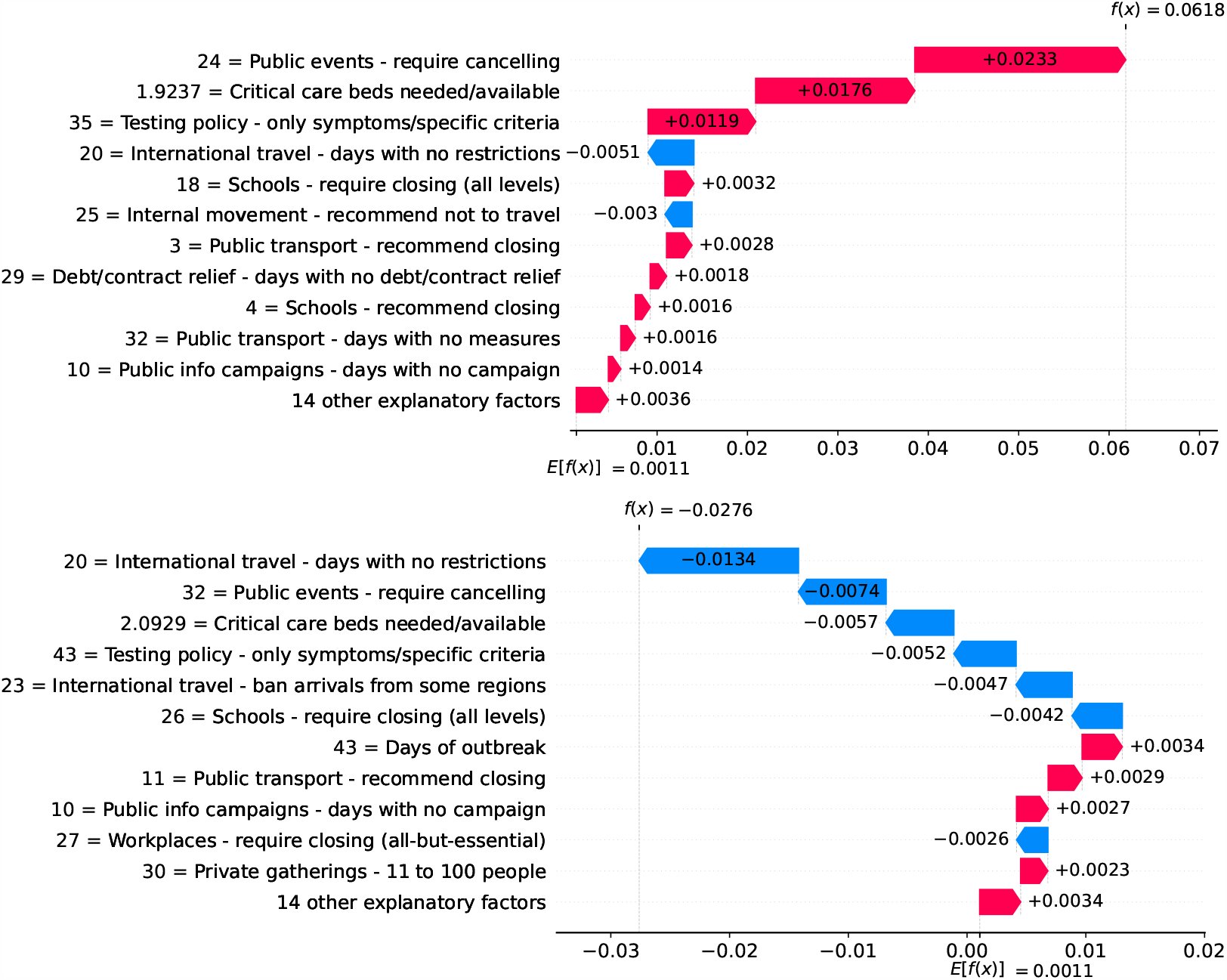
(Color online) Waterfall plots for Netherlands. Factors driving acceleration on the 20^*th*^ (top) and 26^*th*^ day (bottom).

Figure 8 shows observed and predicted curves for the UK. The peak of acceleration occurred on the 23^*rd*^ day after the first death, while the peak of velocity occurred on the 37^*th*^ day. Figure 9 shows the main factors explaining acceleration on the 30^*th*^ and 37^*th*^ days. Few days after the peak of acceleration, measures such as closing schools/universities and self-isolation, started to decrease acceleration. On the peak of velocity, other measures started to become effective. Cancelling public events seems to be an important measure, which still needs additional days to finally become effective.

**Fig 8.**
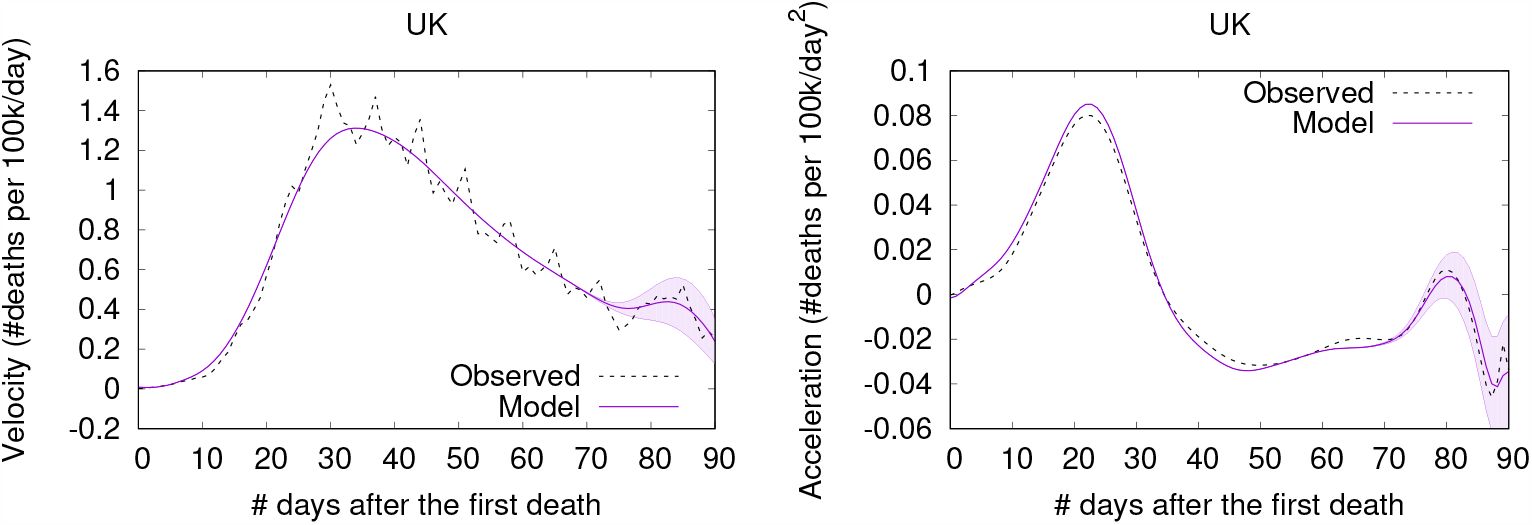
(Color online) Projections for the UK.

**Fig 9.**
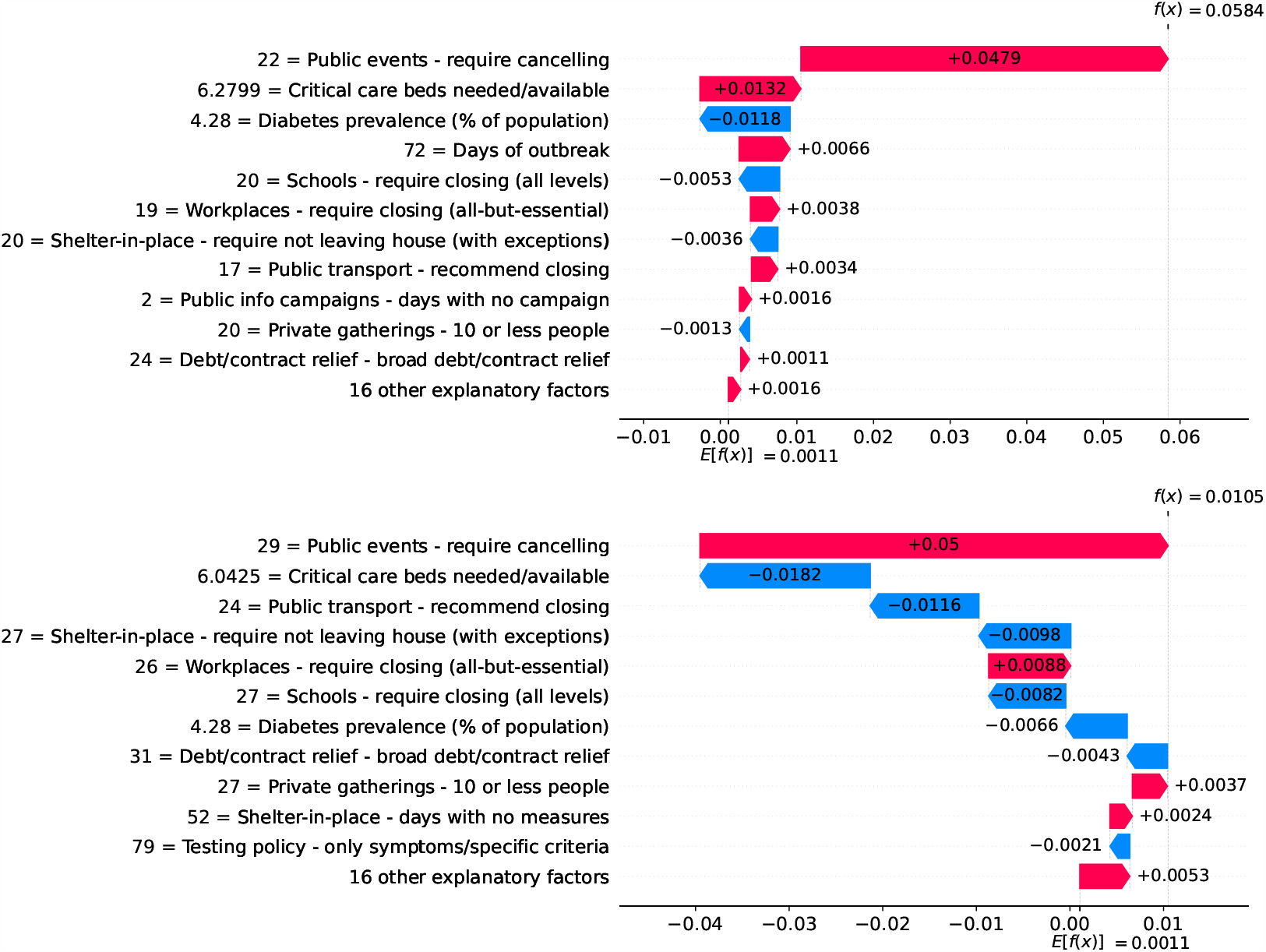
(Color online) Waterfall plots for the UK. Factors driving acceleration on the 30^*th*^ (top) and 37^*th*^ day (bottom).

Countries have adopted different measures, resulting in shorter or longer periods between the peaks of acceleration and velocity. In Netherlands this period lasted for only nine days, while in the UK this period lasted for 14 days.

### Germany - The importance of a robust health system

Figure 10 shows observed and predicted curves for Germany. The curves point to two days of particular interest: the 20^*th*^ day after the first death (which marks the peak of acceleration), and the 39^*th*^ day (which marks the peak of velocity).

**Fig 10.**
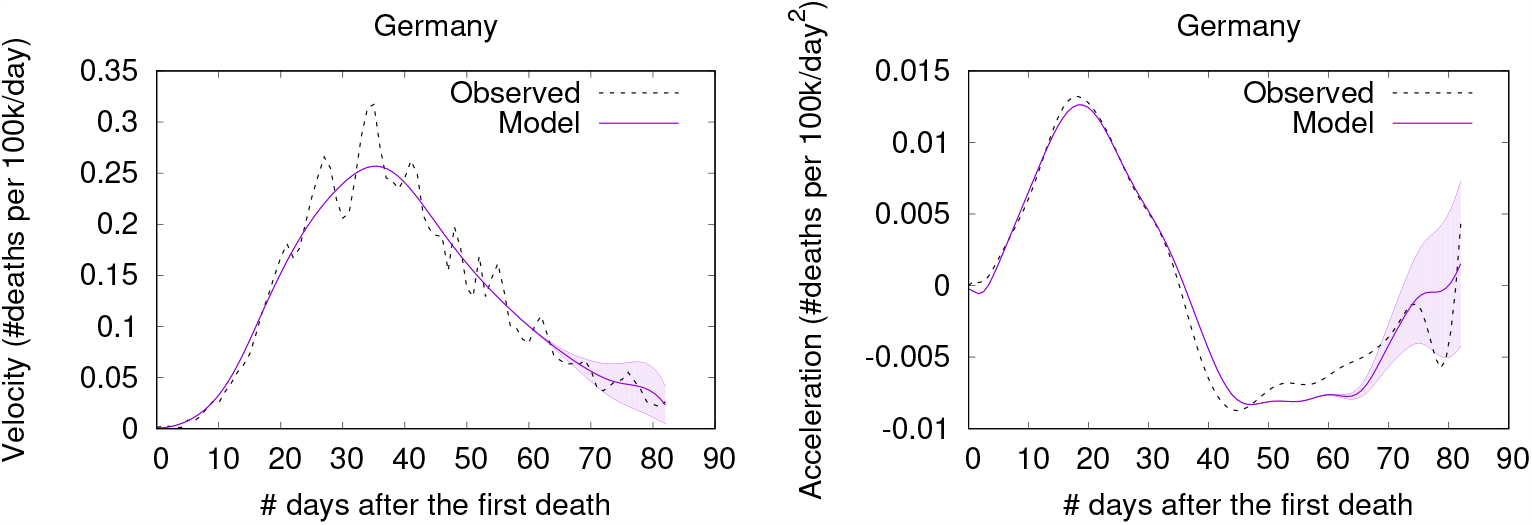
(Color online) Projections for Germany.

Figure 11 shows the factors driving velocity and acceleration are shown on the 20^*th*^ day. Acceleration is mainly explained by the lack and the recency of countermeasures. For instance, no public transport measures were taken for 67 days, and this is the main factor increasing acceleration at this stage of the outbreak. A related factor is the small decrease of visitors in transit stations (−10%), which also increases acceleration.

**Fig 11.**
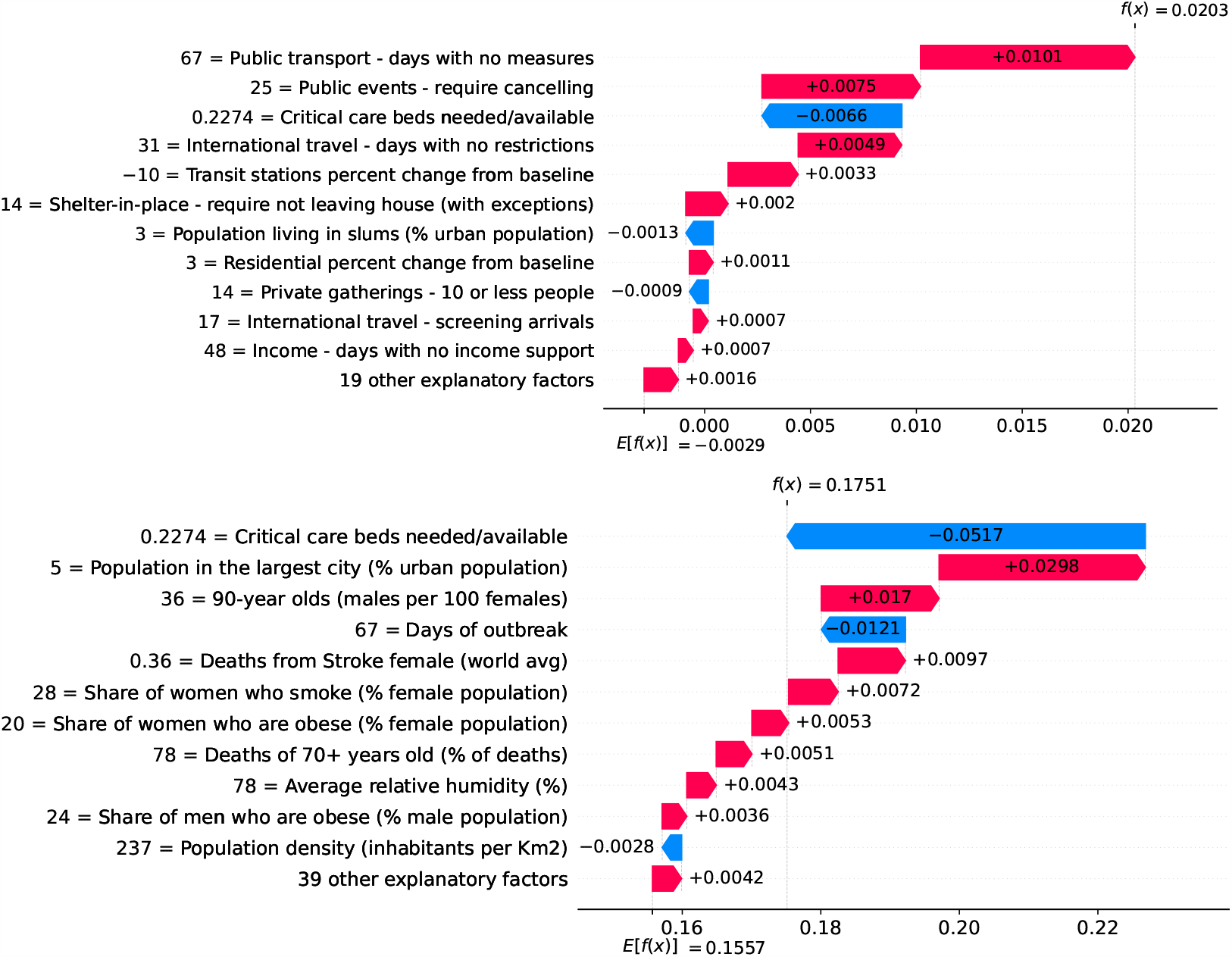
(Color online) Waterfall plots for Germany on the peak of acceleration on the 20^*th*^ day. Top - Factors driving acceleration. Bottom - Factors driving velocity.

Since the 20^*th*^ day marks the peak of acceleration, it is not expected to find many factors contributing to decrease acceleration. In fact, there are only three factors worth mentioning: the small proportion of urban population living in slums, the restriction on private gatherings, and most importantly, the high availability of critical care beds, indicating a robust system even during the peak of acceleration. As expected, many factors are contributing to increase velocity on the peak of acceleration. These factors are shown in the bottom of Figure 11, including the distribution of urban population, male/female proportion on different ages, and risk factors such as obesity and smoking. On the other side, the high availability of critical care beds is the main factor contributing to decrease velocity. This points, again, to the importance of a prepared and robust health system in order to reduce death toll.

Figure 12 shows that on the 39^*th*^ day acceleration became negative. Thus, many factors are contributing to decrease acceleration. Stringent measures (such as school closures, workplace closures and travel bans) were adopted for a time which is now sufficient to show benefits. Further, the high availability of critical care beds keeps contributing to decrease acceleration. The 39^*th*^ day also marks the peak of velocity, and factors contributing to increase velocity include typical comorbidities (i.e., stroke) and risk factors such as obesity. The only relevant factor contributing to decrease velocity is the high availability of critical care beds. Ensuring sufficient physical infrastructure and resources, even on the peak of velocity, is crucial for dealing with the outbreak.

**Fig 12.**
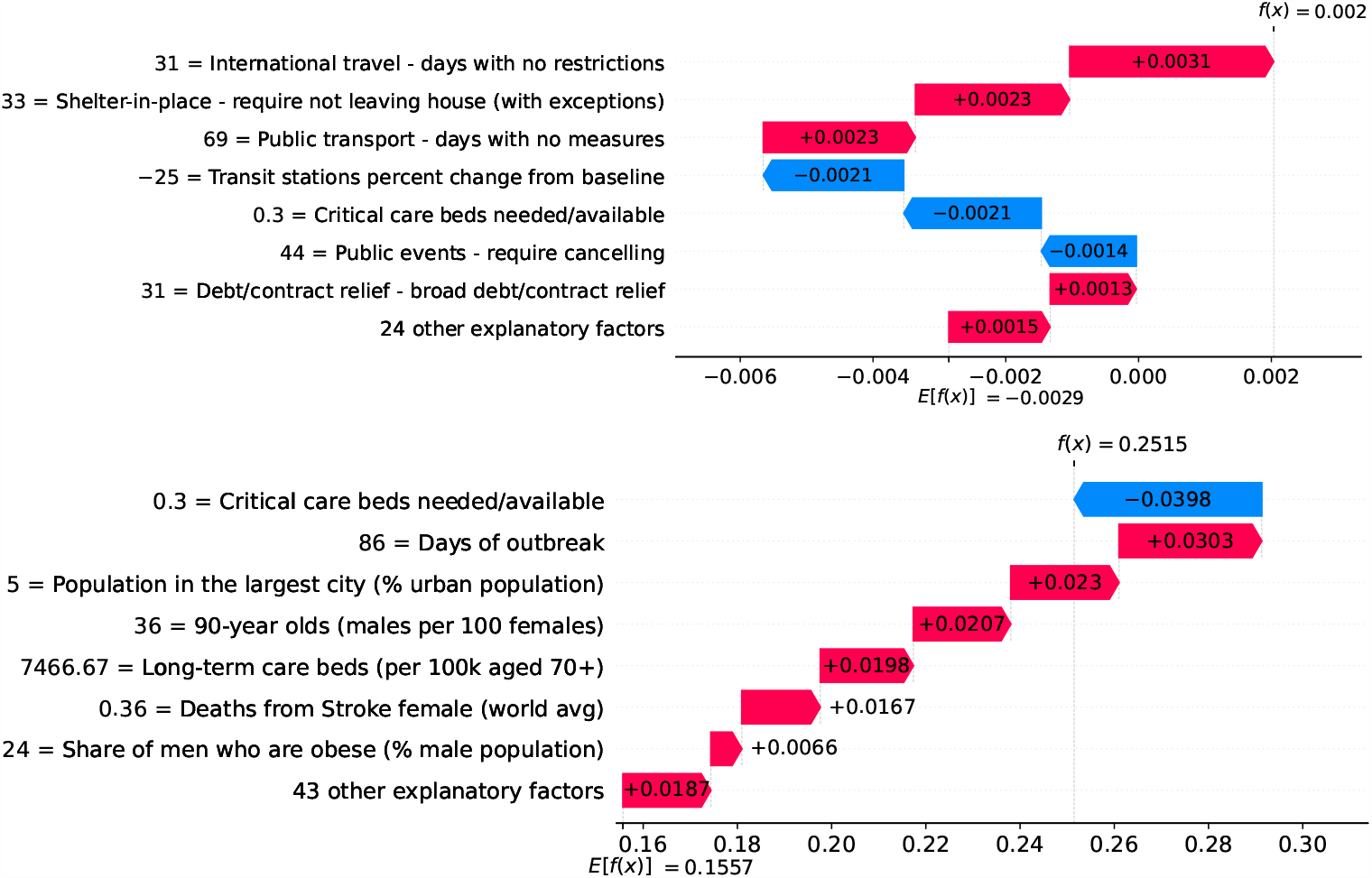
(Color online) Waterfall plots for Germany on the peak of velocity on the 39^*th*^ day. Top - Factors driving acceleration. Bottom - Factors driving velocity.

### Brazil - Urban slums are potential incubators of COVID-19

Figure 13 shows observed and predicted curves for Brazil. A particular period of the outbreak, starting on the 31^*st*^ day and continuing until the 45^*th*^ day, was marked by a consistent increase in acceleration. The drastic increase in acceleration caused the health system to collapse, as critical care beds needed became greater than beds available. Next we discuss the main factors driving acceleration during this period.

**Fig 13.**
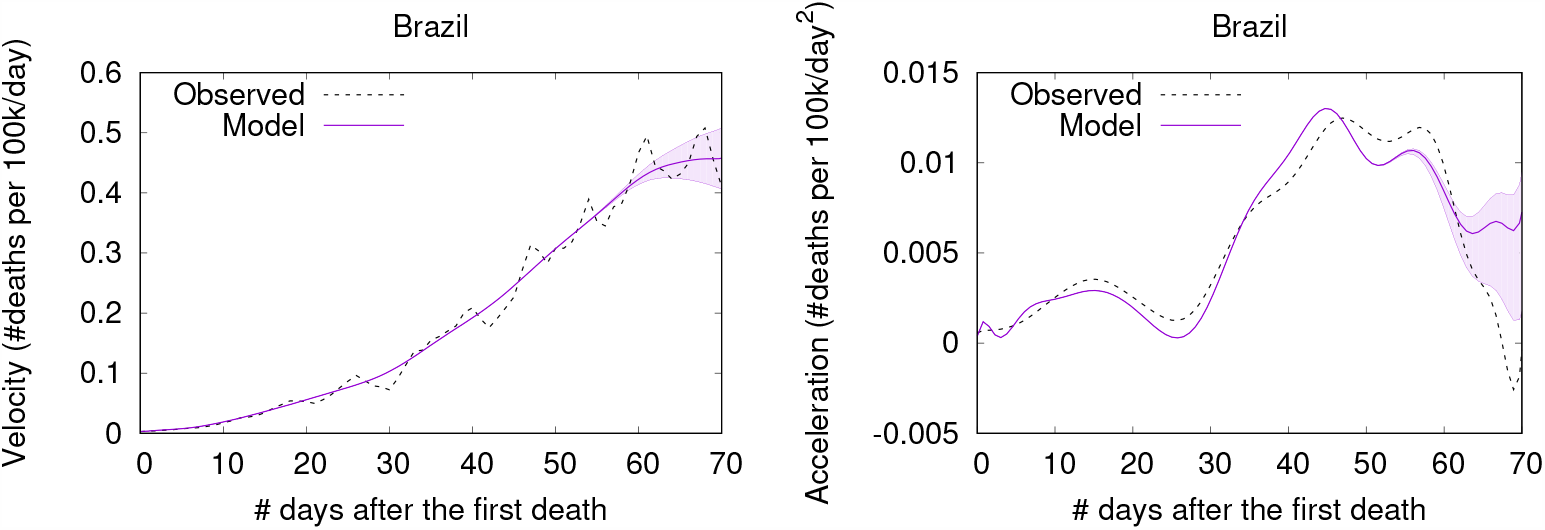
(Color online) Projections for Brazil.

Figure 14 shows the main factors driving acceleration in Brazil from the 31^*st*^ to the 45^*th*^ day after the first death. Clearly, the high proportion of urban population living in slums was a major factor increasing acceleration on this period. Urban slums are characterized by low-quality housing, limited basic services and poor sanitation. This makes simple lifesaving practices like hand-washing a challenge. Further, households in urban slums are typically +10 times denser than neighboring areas of the same city. The largest urban slum in Brazil, for instance, has a density of 39,000 inhabitants per km^2^, while the country’s population density is only 25 inhabitants per km^2^. Finally, many people in slums need to work normally during the outbreak, as they often have no savings. They use public transportation over long distances, and their service occupations usually require close contact with others, making them vectors for the virus and accelerating the spread of infection.

**Fig 14.**
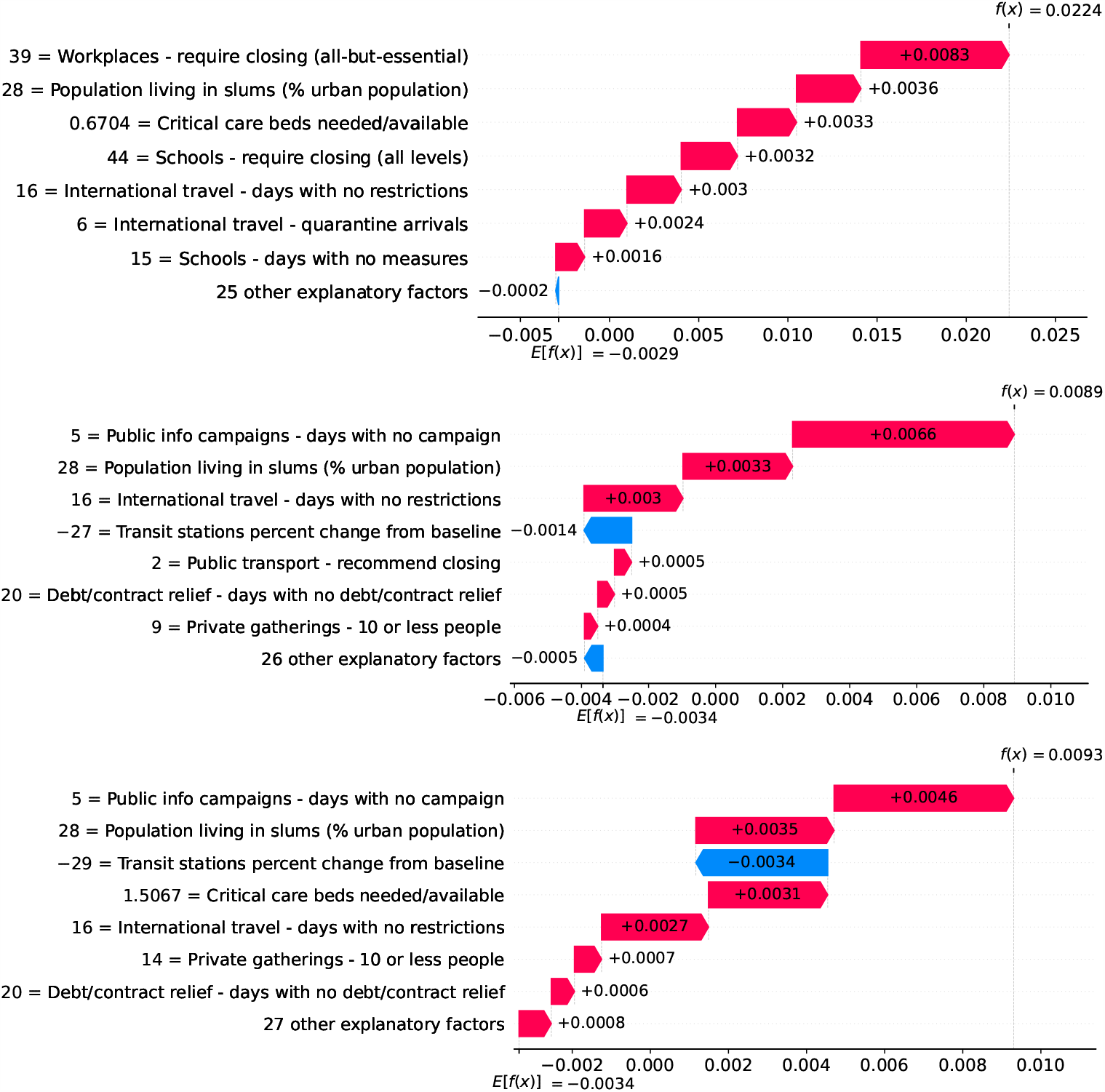
(Color online) Waterfall plots showing factors driving acceleration on different days in Brazil. From top to bottom: 31^*st*^, 40^*th*^, and 45^*th*^ day after the first death.

### Belgium - The risk with long-term care and nursing institutions

A high proportion of long-term care facilities have reported COVID-19 outbreaks, with high rates of morbidity and case fatality in residents. Deaths in Belgium’s long-term care institutions account for one third of the COVID-19 death toll in the country, while in Switzerland, residents of long-term care institutions account for more than half of deaths due to COVID-19 [11]. Despite measures promoting social distancing, long-term care residents need care from staff who inevitably might transmit the virus. Infected asymptomatic staff members ravaged the long-term care institutions whose residents are older adults often with underlying chronic medical conditions [12].

Figure 15 shows observed and predicted curves for Belgium. The curves point to two days of particular interest: the 21^*st*^ day after the first death, which marks the peak of acceleration, and the 30^*th*^ day, which marks the peak of velocity. These days are further inspected in Figure 16, which shows the main factors driving velocity. Indeed, the high number of long-term care beds in Belgium is among the factors contributing to increase velocity on both peaks.

**Fig 15.**
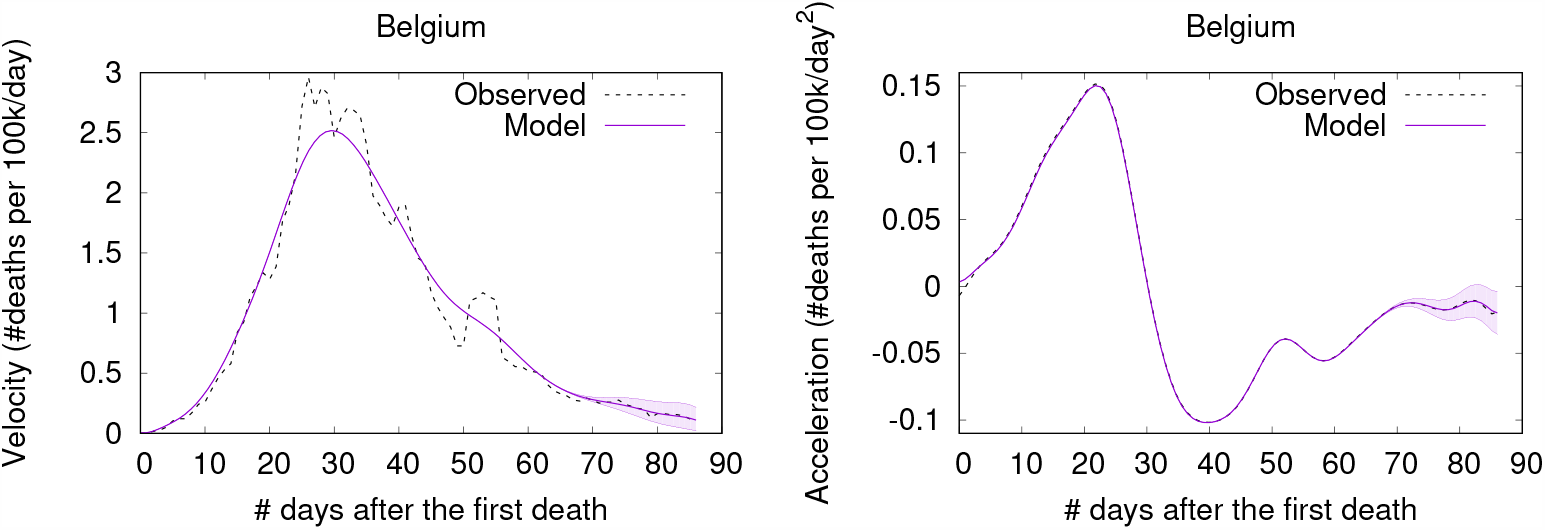
(Color online) Projections for Belgium.

**Fig 16.**
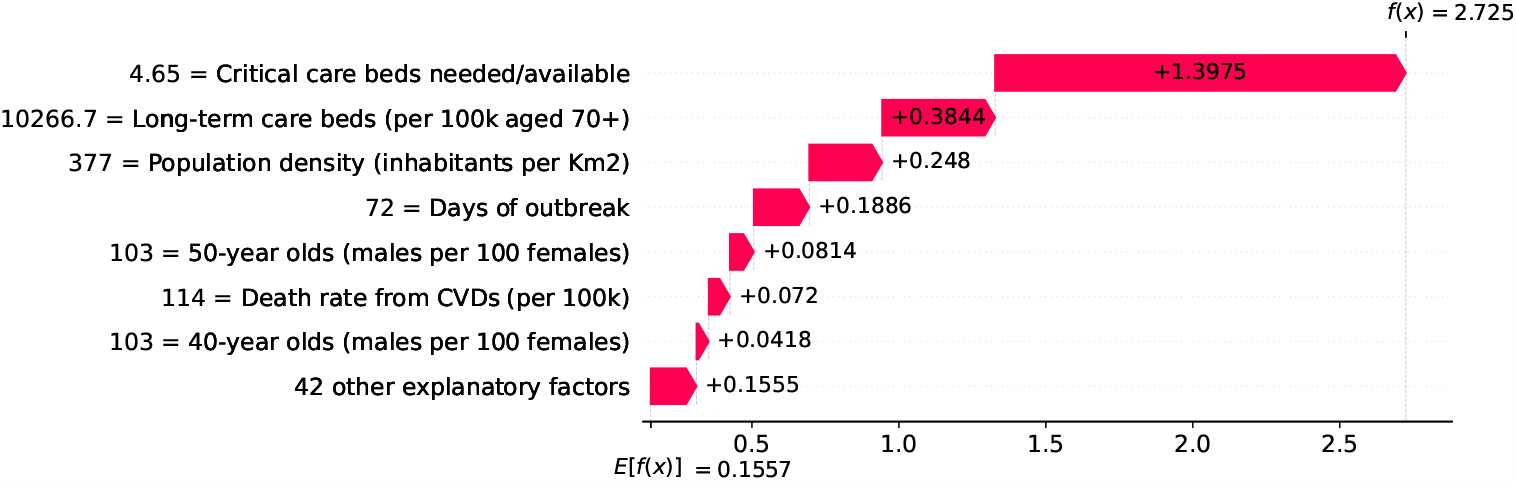
(Color online) Waterfall plots for Belgium. Factors driving velocity on the peak of velocity.

### Brazil and USA - The persistent impact of income inequality

While some factors gain importance only during specific periods of the outbreak, income inequality shows high impact on velocity during the entire outbreak. Income inequality is assessed with the GINI Index [13], a standard measure which varies from 0 (total equality) to 100 (total inequality). Inequality has many undesirable effects including significant negative impacts on public health [14], and our models reveal that countries with greater income inequality are more likely to face an increased COVID-19 death toll pace. This is the case of the two most affected countries, Brazil and the USA, two countries that suffer greatly from high income inequality.

Figures 17 and 18 show the main factors driving velocity on Brazil and USA. Inequality is among the major factors contributing to increase velocity during the entire outbreak. In some specific days, inequality is the most important factor.

**Fig 17.**
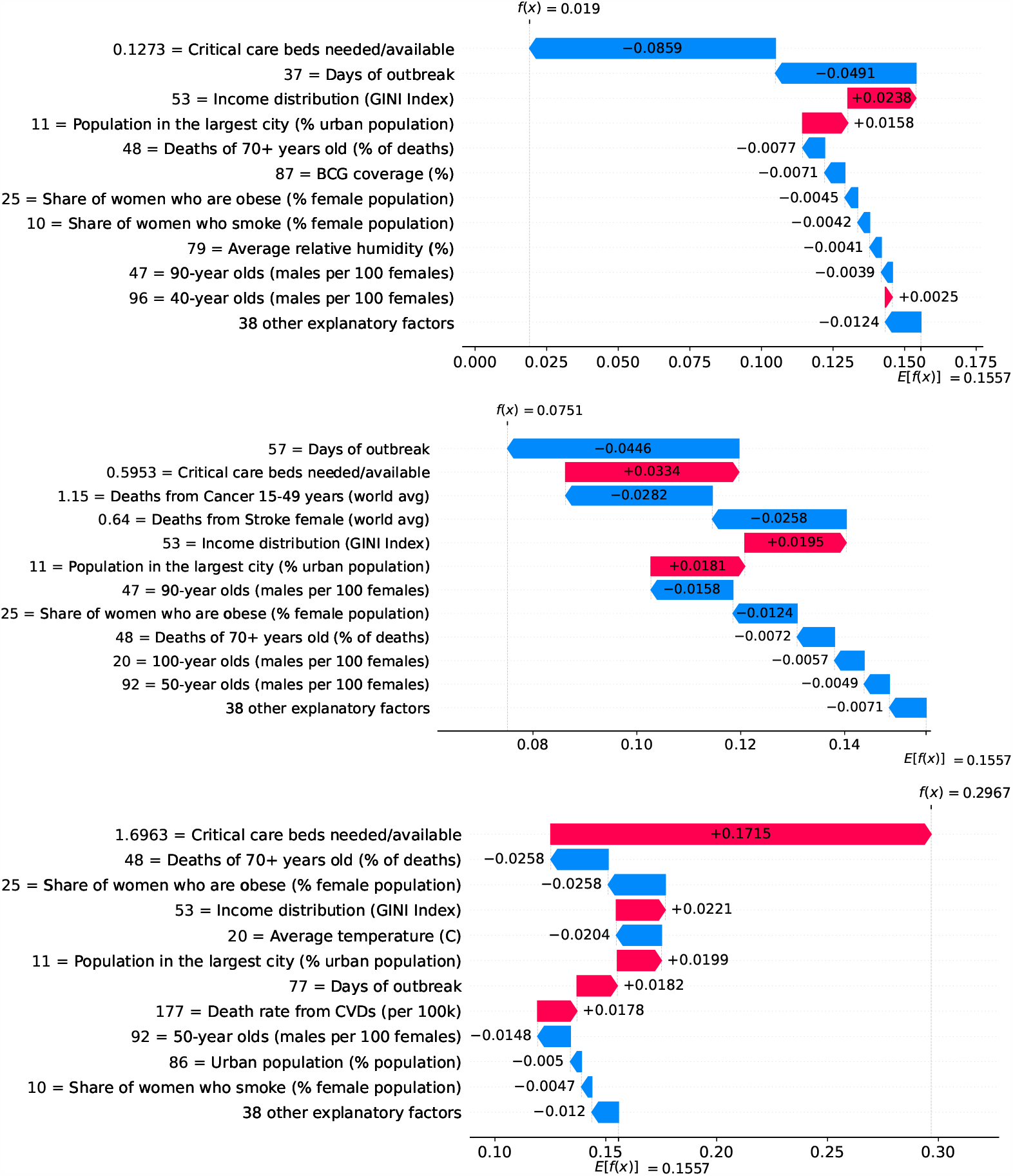
(Color online) Waterfall plots showing factors driving velocity throughout the outbreak in Brazil. From top to bottom: 10^*th*^, 30^*th*^, and 50^*th*^ day after the first death.

**Fig 18.**
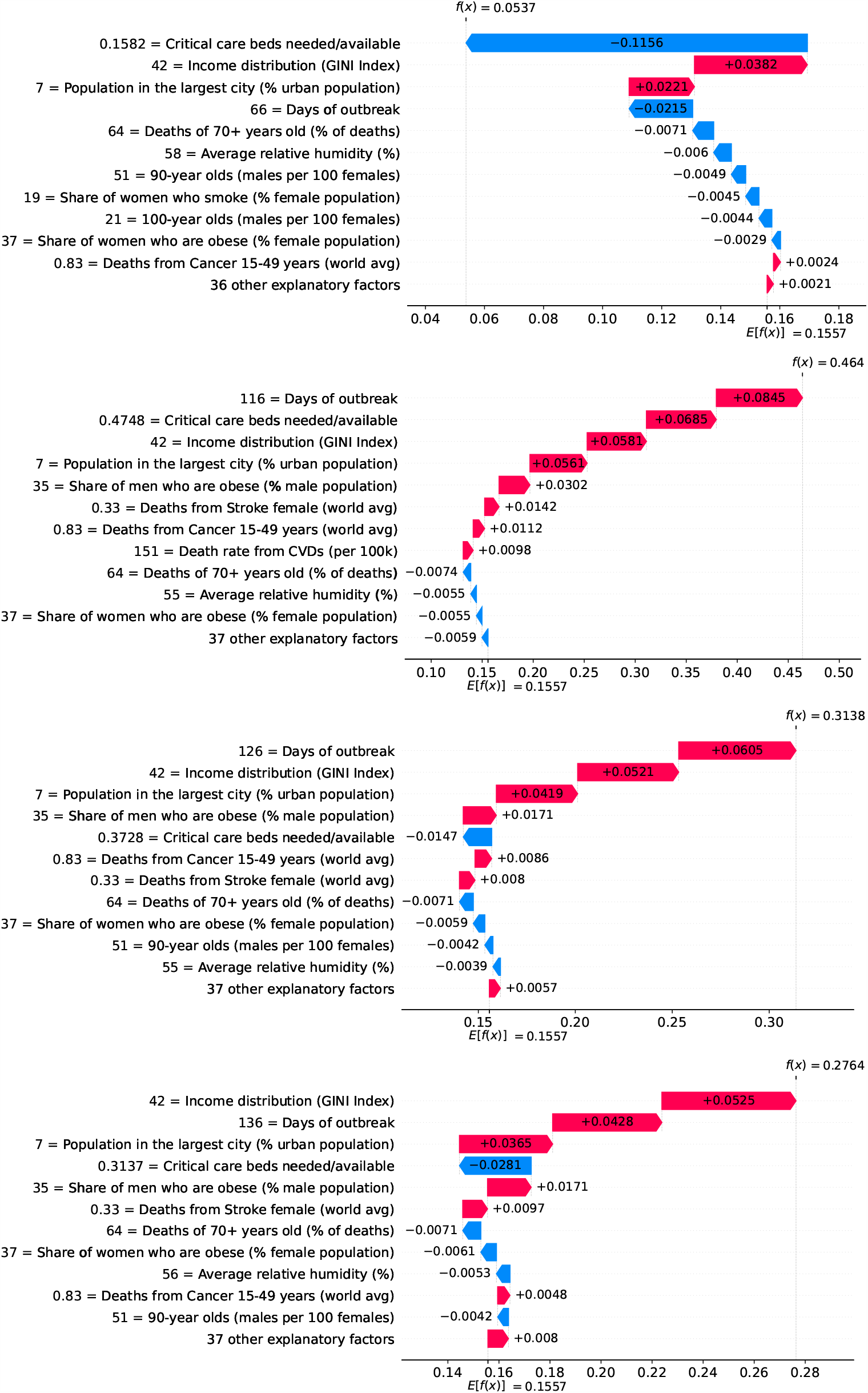
(Color online) Waterfall plots showing factors driving velocity throughout the outbreak in the USA. Top to bottom: 20^*th*^, 60^*th*^, 80^*th*^, and 90^*th*^ day after the first death.

### Global overview on the importance of factors and measures

So far we have discussed how different factors impact velocity and acceleration on specific country-level models. To get a global overview on the impact that different factors have on velocity, we plotted the contribution of every factor for every country. We selected three particular days to analyze: the peak of acceleration, the peak of velocity, and June 5th 2020 as it is the last day in our data. The resulting summary plot has five characteristics:

- Each row corresponds to a factor and has as many dots as countries.
- A dot represents the value of the corresponding factor for a country.
- Red dots indicate that the factor assumes a high value for the corresponding country. Likewise, blue dots indicate that the factor assumes a low value.
- The vertical line shows whether the impact of a factor increased the prediction (i.e., the dot is on the right size) or decreased it (i.e., the dot is on the left side).
- Factors impacting most the model prediction appear on the top of the plot.

Figure 19 shows how the different factors are explaining velocity at the peak of acceleration on different countries. Next we discuss the role of some of these factors.

**Fig 19.**
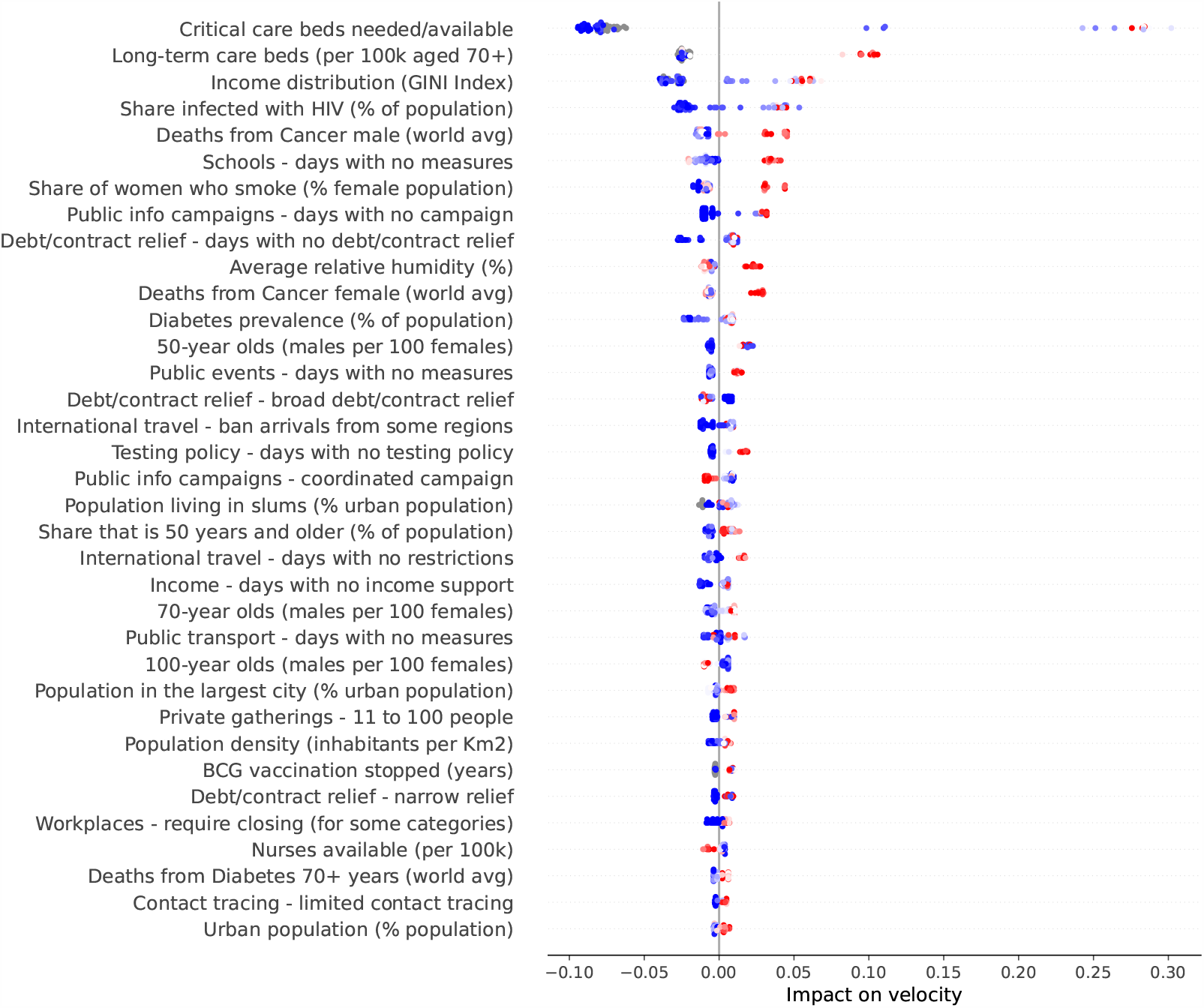
(Color online) Summary plot showing how each factor contributes to increase/decrease velocity at the peak of acceleration.

As expected, the most important factor is the demand for critical care beds. Clearly, countries showing high demands at the peak of acceleration (i.e., red dots) have experienced a large increase in velocity due to this factor. Little demands, on the other hand, usually contribute to decrease velocity. Other generally important factors include income distribution, number of beds in long-term care institutions, smoking prevalence among females, share of population infected with HIV, and relative number of cancer-related deaths among the elderly population. High values of these factors contribute to increase velocity.

It is worth mentioning that while we are evaluating each factor in isolation, their contributions to velocity are calculated by taking into account the complex interplay among all them. Take for instance the role of relative humidity - low relative humidity contributes to decrease velocity, while high relative humidity usually contributes to increase velocity. There are some exceptions, however, where high relative humidity contributes to decrease velocity. In order to better understand these exceptions, we extracted a heatmap from our working data showing the relationship between temperature and relative humidity, and check how their interplay ultimately impacts velocity.

Figure 20 presents a heatmap showing the relationship between temperature and relative humidity. Clearly, temperatures higher than *≈*25°C are strongly associated with low velocities, despite the high values of relative humidity.^3^

**Fig 20.**
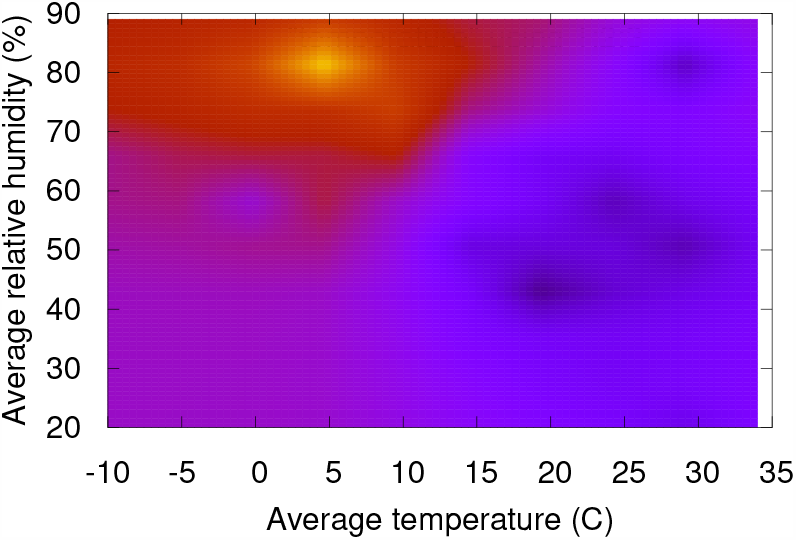
(Color online) The relationship between temperature, relative humidity and velocity. Lighter regions are associated with higher velocities.

The lack of countermeasures to slow the spread of the virus is reflected in several factors occupying the middle of the heatmap. As expected, the more days without a measure the higher is the impact on increasing velocity. A particular important measure at this stage of the outbreak seems to be the adoption of coordinated public information campaign due to the need for essential information and advice right on early stages of the outbreak. Clearly, the more days with coordinated public campaigns the more velocity decreases. However, at this stage of the outbreak, countermeasures are among the factors impacting the least on velocity.

Other factors that are worth mentioning include the number of years without BCG vaccination and share of one-person households in the country. Both factors show marginal impact on velocity at the peak of acceleration and are among the ones showing small impact on velocity, but they gain importance on latter stages of the outbreak.

Figure 21 shows how the different factors impact velocity at the peak of velocity on different countries. Top factors remain the same as the ones at the peak of acceleration, namely: demand for critical care beds, number of long-term care beds in institutions, and income distribution. Other factors, such as number of years without BCG vaccination gained substantial importance. Specifically, some countries seem to benefit from an active national vaccination program. As recently reported in [16], the BCG vaccine has beneficial non-specific effects on the immune system that protect against a wide range of infections, and randomized controlled trials have provided evidence that the BCG vaccine’s immunomodulatory properties can protect against respiratory infections.

**Fig 21.**
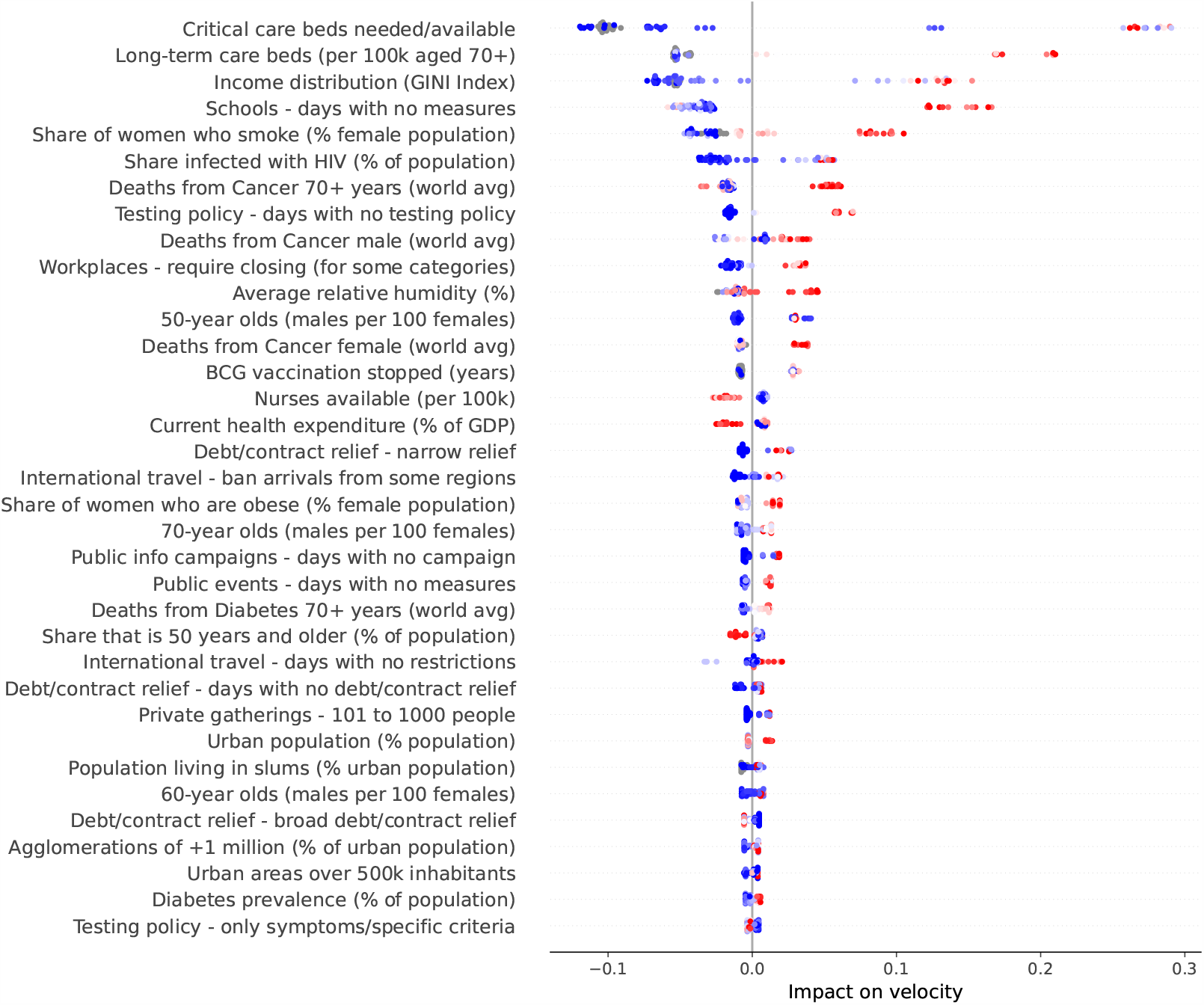
(Color online) Summary plot showing how each factor contributes to increase/decrease velocity at the peak of velocity.

Two factors that also gained substantial importance are the availability of nurses and the percentage of GDP expended on the health system. Specifically, velocity decreases as more nurses are available. Similarly, velocity decreases as more is spent on the health system.

Finally, the more days adopting less stringent measures (i.e., closing workplaces only for some categories, narrow debt relief and banning arrivals only from some regions), the more velocity increases. The opposite trend is observed with more stringent measures (i.e., broad contract relief), that is, the more days adopting these measures, the more velocity decreases.

Figure 22 shows how the different factors have impacted velocity on June 5th 2020, the last day in our working data. Interestingly, income distribution became the top factor impacting velocity. The number of nurses available, the age working dependency ratio and the share of elderly people also gained importance, and are now among the factors impacting velocity most.

**Fig 22.**
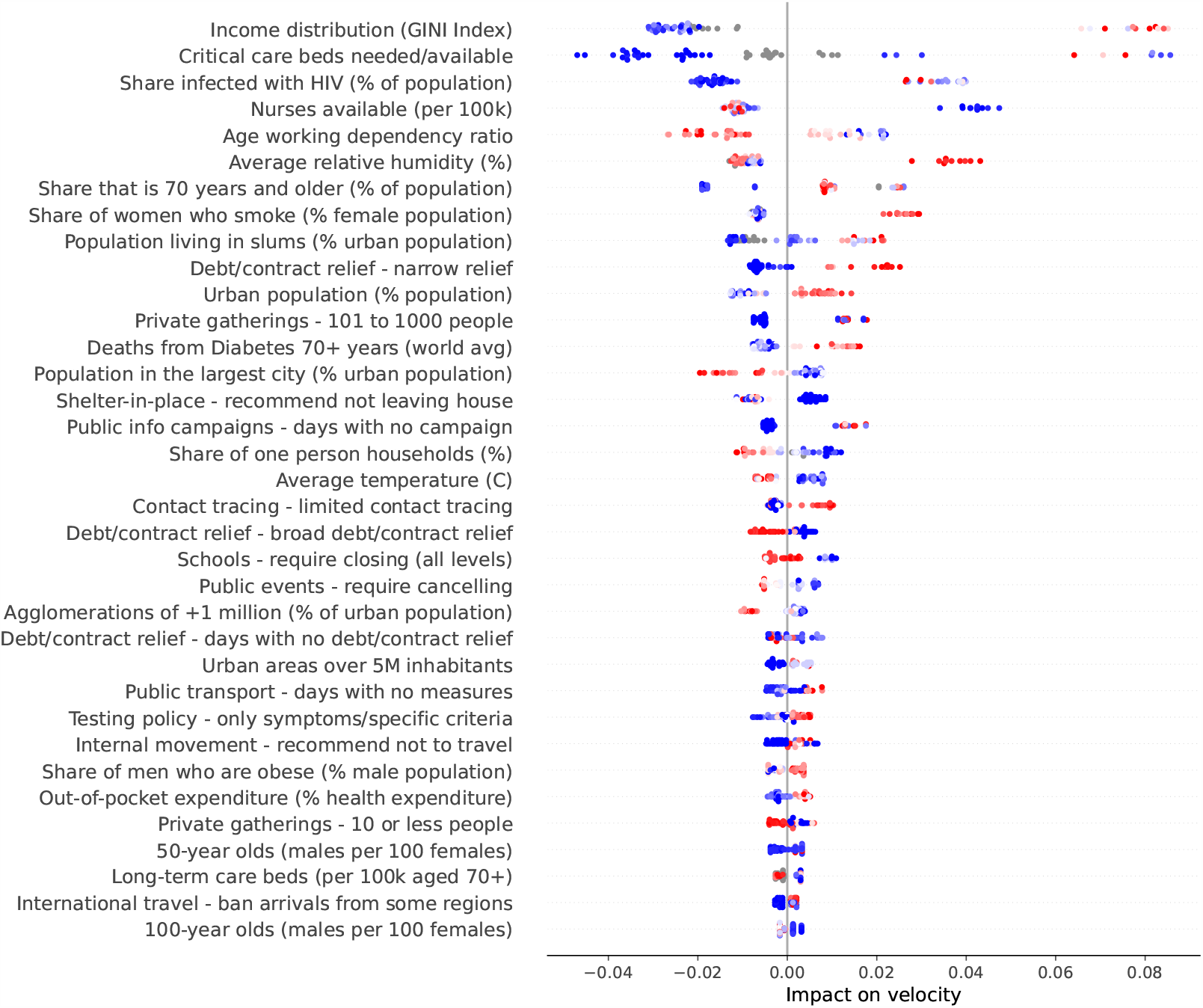
(Color online) Summary plot showing how each factor contributes to increase/decrease velocity on June 5th 2020.

Two factors are involved in an interesting contrast: the share of one-person households and the population living in slums. Both factors have similar importance at this stage of the outbreak, but they show opposite trends. One person households contribute to decrease velocity, while population living in slums contribute to increase velocity. People living together are clearly more exposed to getting infected because they share space. Larger households, which are a characteristic of urban slums, pose greater risk of infection during the outbreak because they can contribute to the rapid spread of the virus.

Finally, at this stage of the outbreak, it becomes clearer which countermeasures were effective. We consider as effective a measure which contributes to decrease velocity as it is adopted for more days. Thus, effective measures include the recommendation of not leaving house, the adoption of a broad debt relief, closing schools, limiting private gatherings to less than 10 people, and cancelling public events.

### Predicting counterfactuals

Our counterfactual scenarios are built by performing hypothetical variations on countermeasures that are being actually adopted in a country [17]. By performing these variations, the overall stringency level of the country is increased or decreased accordingly, following Equation 1. As a result, for each day *d* we have many different scenarios based on hypothetical variations of the countermeasures being adopted. As an example, suppose a country where the countermeasure “Ban arrivals from some regions” was adopted on day *d*. In this case, we create a hypothetical scenario for day *d* in which the country would be adopting countermeasure “Screening arrivals” instead. In this case, the overall stringency level decreases. Alternatively, we may create a hypothetical scenario in which the country would be adopting “Ban arrivals from all regions” on day *d*, and in this case the overall stringency level is increased. Therefore, we have different hypothetical scenarios for each day of the outbreak.

Now, let *ν*_*d*−1_ be the observed velocity on day *d*−1. Clearly, *ν*_*d*−1_ is the fact obtained as the result of adopting a specific set of countermeasures in the country. Lets assume we have built *n* hypothetical scenarios for each day of the outbreak, from which we use our models to predict the corresponding accelerations for day *d*. This results in a set of *n* predicted accelerations for day 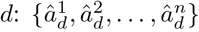, one prediction for each counterfactual scenario. The next step is to obtain the counterfactual velocities for the next day: 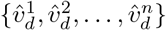, where each 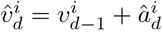. Notice that each counterfactual scenario *i* is associated with a different stringency level. The death toll associated with a counterfactual scenario *i* is simply given as the sum of the corresponding counterfactual velocities over all days of outbreak, that is: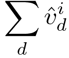.

Finally, we can easily calculate the difference between the stringency level of a counterfactual scenario and the actual/observed stringency level in the country. A positive difference means that the corresponding counterfactual scenario is more stringent than the actual scenario. On the other hand, a negative difference reveals that the counterfactual scenario is less stringent than the actual scenario. Likewise, we calculate the difference between the actual death toll and the counterfactuals, and this difference corresponds to our expectation of the counterfactual effects. Since we have considered many counterfactual scenarios, we can simply calculate the interval of death toll change, that is, a specific counterfactual scenario may lead to less change than others, even if they have the same stringency level.

Figure 23 shows the counterfactual effects of hypothetical countermeasures in the death toll for different countries. According to our results, Belgium would have experienced a drop of up to 9% on death toll if the adopted countermeasures were more stringent. On the other hand, death toll would have increased 8% with the adoption of less stringent countermeasures. We provide the same analysis for UK, Sweden and USA.

**Fig 23.**
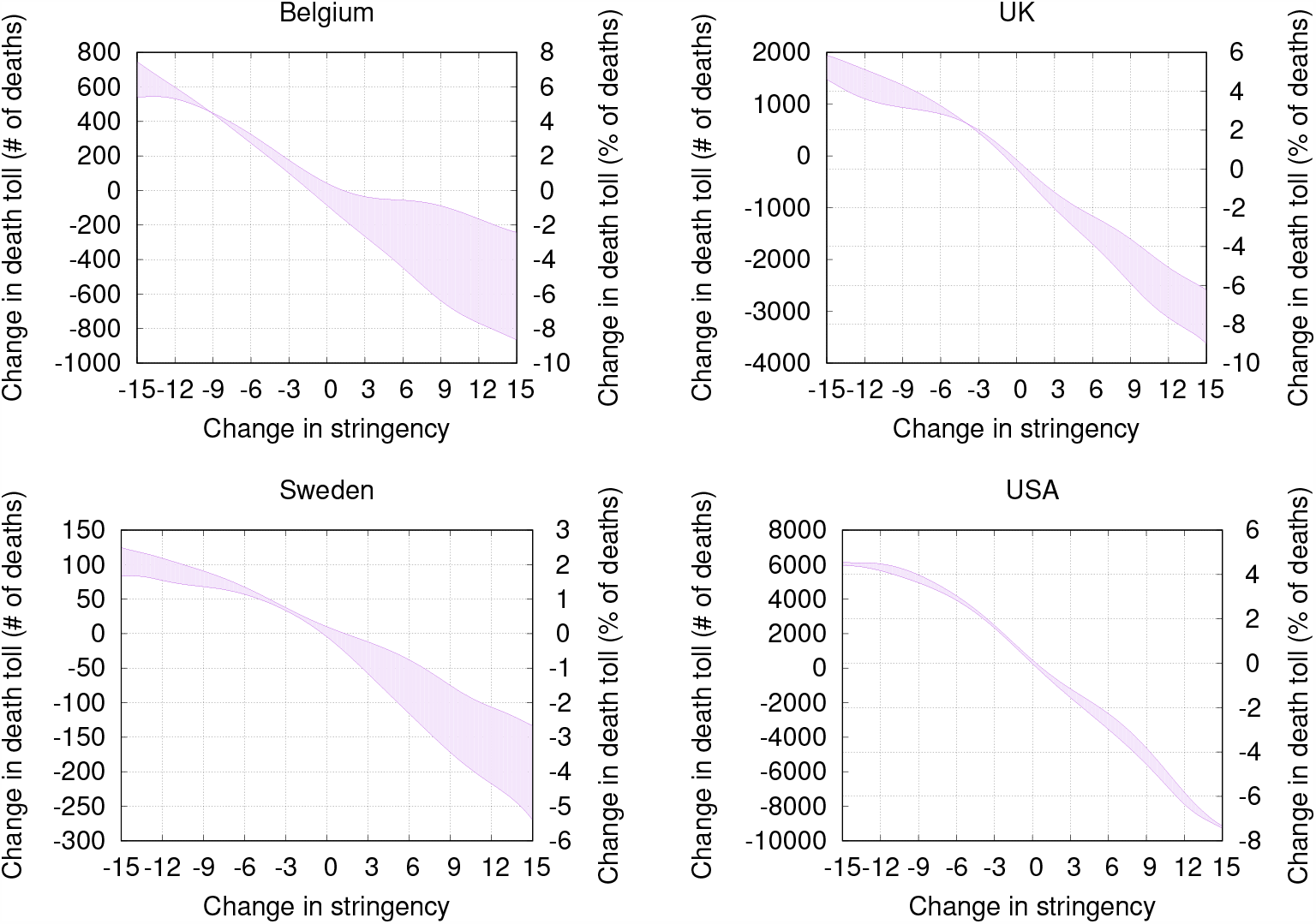
(Color online) Counterfactual death toll predictions for different countries.

An interesting additional dimension for our counterfactual analysis would be to predict the corresponding effects on the economy. Unfortunately, we do not have the necessary data to include the economics dimension. However, in the study [18], the authors used geographic variation in mortality during the 1918 Flu Pandemic in the USA and found that more exposed areas experience a sharp and persistent decline in economic activity. On the contrary, cities that intervened earlier and more aggressively experienced a relative increase in real economic activity after the pandemic subsided. Altogether, their findings suggest that pandemics can have substantial economic costs, and non-pharmaceutical interventions can lead to both better economic outcomes and lower mortality rates.

## Conclusion

This study shows how useful concepts in model explainability and counterfactuals can be used to facilitate reasoning about different hypotheses regarding the ongoing COVID-19 pandemic. For this, we built intuitive country-level COVID-19 motion models described in terms of death toll velocity and acceleration, which are defined as the first and second order derivatives, respectively, of the COVID-19 death toll in respect to time. The study considered a multitude of factors, and we show that both velocity and acceleration can be approximated with proper combinations of these factors. Model explanation techniques reveal the main factors driving velocity and acceleration. In general, more intricate factors such as urbanization problems, typical comorbidities, and social inequality issues are among the ones better explaining velocity. Acceleration, on the other hand, is explained by the effects of public health measures being adopted, or the lack of them. These explanations enable data-driven narratives that may help epidemiologists and policymakers to understand factors driving the outbreak, as well as to monitor the impact of public health measures on the spread of the disease. We also provide counterfactual predictions in order to estimate what is likely to happen as a result of an action. As with all approaches to causal inference on non-experimental data, strong assumptions were required [19]. Specifically, we assumed an independence between countries in the sense that increasing/decreasing the overall stringency level of a country does not change the factors and the death toll observed in other countries. Still, our predictions allow the assessment of the impact of countermeasures by means of “what-if” simulation scenarios.

## Data Availability

All data used in our study is freely available, and the data sources are properly stated in the manuscript.

## Acknowledgments

We thank the support given by the Project: Models, Algorithms and Systems for the Web (grant by FAPEMIG / PRONEX / MASWeb APQ-01400-14) and authors’ individual grants and scholarships from CNPq. We also thank KUNUMI for the financial support of the Artificial Intelligence Laboratory of the Department of Computer Science at UFMG, where this research was carried out.

## Footnotes

1 https://cloud.google.com/maps-platform

2 Please, visit https://covid-19.kunumi.com for a live instance of our analysis tool with a larger set of countries.

3 Interestingly, authors in [15] have reported that the relationship between the annual average of temperature compensation and COVID-19 confirmed cases was approximately linear in the range of less than 25.8°C, which became flat above 25.8°C.

